# The BAF complex inhibitor pyrimethamine reverses HIV-1 latency in people with HIV-1 on antiretroviral therapy

**DOI:** 10.1101/2022.09.23.22280188

**Authors:** H.A.B. Prins, R. Crespo, C. Lungu, S. Rao, L. Li, R.J. Overmars, G. Papageorgiou, Y.M. Mueller, T. Hossain, T.W. Kan, B.J.A. Rijnders, H.I. Bax, E.C.M. van Gorp, J.L. Nouwen, T.E.M.S. de Vries-Sluijs, C.A.M. Schurink, M. de Mendonça Melo, E. van Nood, A. Colbers, D. Burger, R-J. Palstra, J.J.A. van Kampen, D.A.M.C. van de Vijver, T. Mesplède, P.D. Katsikis, R.A. Gruters, B.C.P. Koch, A. Verbon, T. Mahmoudi, C. Rokx

## Abstract

A major barrier towards HIV-1 cure is the presence of a replication-competent latent reservoir that, upon treatment cessation, can spark viral rebound leading to disease progression. Pharmacological reactivation of the latent HIV-1 reservoir with Latency reversing agents (LRAs) is a first step toward triggering reservoir decay. Inhibitors of the BAF-complex, a key repressor of HIV-1 transcription were identified to act as LRAs, and enhanced the effect of other LRAs such as histone deacetylase inhibitors ex-vivo. We repurposed the licensed drug pyrimethamine as a BAF-inhibitor to investigate its *in vivo* impact on the HIV-1 reservoir of people living with HIV-1 (PLWH). Twenty eight PLWH on suppressive antiviral therapy were randomized in a 1:1:1:1 ratio to receive pyrimethamine; high dose valproic acid; both valproic acid and pyrimethamine; or no intervention for 14 days. The primary endpoint was change in HIV-1 reactivation measured as cell associated (CA)HIV-1 RNA at treatment initiation and at the end of treatment. We observed a rapid, modest and significant increase in CAHIV-1 RNA in CD4+T-cells in response to pyrimethamine exposure, which persisted throughout the 14 day treatment, concomitant with induction of BAF target genes as biomarkers of pyrimethamine activity as well as detected plasma pyrimethamine levels. Valproic acid treatment alone did not lead to increase in CAHIV-1 RNA, nor did valproic acid augment the latency reversal effect of pyrimethamine. Despite demonstrated latency reversal, pyrimethamine treatment did not result in a reduction in the size of the inducible reservoir as determined by a tat/rev limiting dilution assay. Serious adverse events were not observed, although physician-directed treatment adjustments occurred, particularly when combining valproic acid with pyrimethamine. These data underline the need for pharmacovigilance in combinatorial clinical strategies and demonstrate that the BAF inhibitor pyrimethamine reverses HIV-1 latency *in vivo* in PLWH, substantiating its potential in advancement in clinical studies to target the proviral reservoir. Clinicaltrials.gov:NCT03525730

**One sentence summary:** This clinical trial shows that the BAF inhibitor pyrimethamine reverses HIV-1 latency in vivo which supports repurposing this drug for cure studies.

## INTRODUCTION

Despite drastically improved treatment prospects brought about by combination antiretroviral therapy (cART), an infection with the human immunodeficiency virus type 1 (HIV-1) remains a chronic condition that, with few exceptions, necessitates lifelong cART. A pool of long-lived CD4+ T cells harbors latent, replication-competent provirus integrated in the genome. This latent HIV-1 reservoir is established in the first weeks after HIV-1 infection and persists stably even after prolonged periods of cART (*1*). Treatment cessation results in rapid plasma viral rebound causing disease progression in people living with HIV-1 (PLWH) (*2, 3*). Therefore, interventions beyond cART are needed towards an HIV-1 cure.

One of the key approaches currently proposed to eradicate the latent HIV-1 reservoir in PLWH is to pharmacologically reverse latency via latency reversing agents (LRAs). Elimination of the re-activated reservoir cells could then be triggered by intrinsic pro-apoptotic pathways or via extracellular immune cell-mediated mechanisms (*4–6*). The ultimate aim is to accomplish a prolonged state of sustainable viral remission without cART in PLWH.

The multitude of LRAs that have been identified through mechanism-based approaches or large-scale screening of compound libraries can be categorized based on distinct molecular and pharmacological targets (*7, 8*), all leading to reactivation of HIV-1 transcription. The LRA classes that influence this process can be classified as de-repressors antagonizing the repressive latent HIV-1 promotor locus, or as activators that directly stimulate viral transcription initiation or elongation. The importance of blocks at the post-transcriptional level have also come to light recently, and may be targetable by LRAs (*9–12*). In contrast to the numbers identified *in vitro*, only a few LRAs have regulatory approval and can currently be used in clinical settings. The deacetylation of histones has been identified and extensively studied as a major mechanism to promote HIV-1 latency (*13–15*), and histone deacetylase inhibitors (HDACis) have become the main evaluated LRA class in HIV-1 cure clinical trials (*16*). The first case series used the HDACi valproic acid and showed an impact on the reservoir of PLWH who also received intensified cART (*17*). However, subsequent larger clinical studies using valproic acid as HDACi without intensified cART did not demonstrate a reduction in the size of the reservoir (*18–22*). Other studies were conducted with clinically approved HDACis that were repurposed as LRA including vorinostat (*23–33*), panobinostat (*34*), and romidepsin (*35–39*) and showed that, in general, HDACis can safely induce HIV-1 transcription at tolerable concentrations in PLWH. However, no HDACi led to a significant reduction of the proviral reservoir when used alone. Studies on the limited number of other LRA classes explored in PLWH have resulted in similar conclusions (*40–43*). Cumulative evidence therefore indicates the need for identification of new LRA classes that potently reactivate HIV-1 and induce reservoir eradication and/or to use combinatorial LRA approaches to reach the potency required to obtain a significant impact on the size of the replication competent reservoir.

Our preclinical studies identified BRG-Brahma Associated Factors (BAF) complex inhibitors (BAFi) as a novel LRA class which we demonstrated enhanced the activity of other LRA classes (*44–46*). The BAF (mammalian SWI/SNF) chromatin remodeling complex is a key repressor of HIV-1 latency that uses ATP hydrolysis to position repressive nucleosome nuc-1 at the HIV-1 long terminal repeat, causing transcriptional silencing of HIV-1 gene expression thereby maintaining HIV-1 latency (*47, 48*). The pharmacological inhibition of the BAF complex leads to de-repression of HIV-1 transcription resulting in latency reversal (*44, 45*). The clinically licensed drug pyrimethamine was subsequently identified as a potent BAFi to reverse HIV-1 latency in primary cell models of latency and in cells obtained from PLWH on cART at tolerable concentrations for humans (*44*). Pyrimethamine also enhanced the activity of other LRAs, including HDACis, in cell lines and primary cell models of HIV-1 latency (*44*). Pyrimethamine is an orally administered, inexpensive antiprotozoan drug widely used in humans, including in people with AIDS, rendering pyrimethamine an attractive candidate for LRA studies (*49–57*).

To study the efficacy of BAF complex inhibition by pyrimethamine to reactivate the HIV-1 reservoirs in PLWH, we conducted a proof-of-concept, randomized controlled clinical trial (LRAs United as a Novel Anti-HIV strategy, LUNA, clinicaltrials.gov:NCT03525730). Apart from a monotherapy arm, we designed a parallel arm where pyrimethamine was combined with an HDACi as partner drug to study an all-oral, affordable, generic combination of LRAs. In contrast to previous studies we chose the HDACi valproic acid at maximized dose, in an acid resistant enteric formulation for better gastrointestinal drug absorption (*18–22*). While previous studies using valproic acid alone did not show a reduction in the reservoir size in PLWH, valproic acid had thus far never been used in combination with other LRAs (*18–22*). We therefore designed the study to examine whether the activity of the individual LRAs can be potentiated *in vivo*. We enrolled participants on suppressive cART and randomized them to 14 days of pyrimethamine (200 mg QD day 1, then 100 mg QD), valproic acid (30 mg/kg/day in 2 equal doses), pyrimethamine and valproic acid, or control with a 4-week follow-up after treatment to study clinical safety and efficacy. Our findings demonstrate that pyrimethamine reactivated HIV-1 *in vivo* in PLWH on suppressive cART, supporting its further development into clinical strategies to target the latent reservoir for reactivation. Despite reversal of latency, pyrimethamine treatment did not lead to a reduction in the size of the inducible reservoir.

## RESULTS

### Clinical characteristics, safety and drug tolerability

Twenty-eight individuals living with HIV-1 on suppressive cART were enrolled and randomized into four arms of seven patients including three interventional arms and one control arm (**Fig. 1**). All participants continued daily cART throughout the study. Participants were men, predominantly white European and on cART for a median of 7.5 years (IQR 5.6-11.7) with a median CD4+ T cell count of 665 cells/μl (IQR 530-820) at inclusion (**Table 1**). Integrase inhibitor based cART regimens were used by 43% of participants, the remainder used non-nucleoside reverse transcriptase inhibitor based cART. Baseline characteristics were comparable between the study arms including the median duration of therapy.

**Fig. 1.**
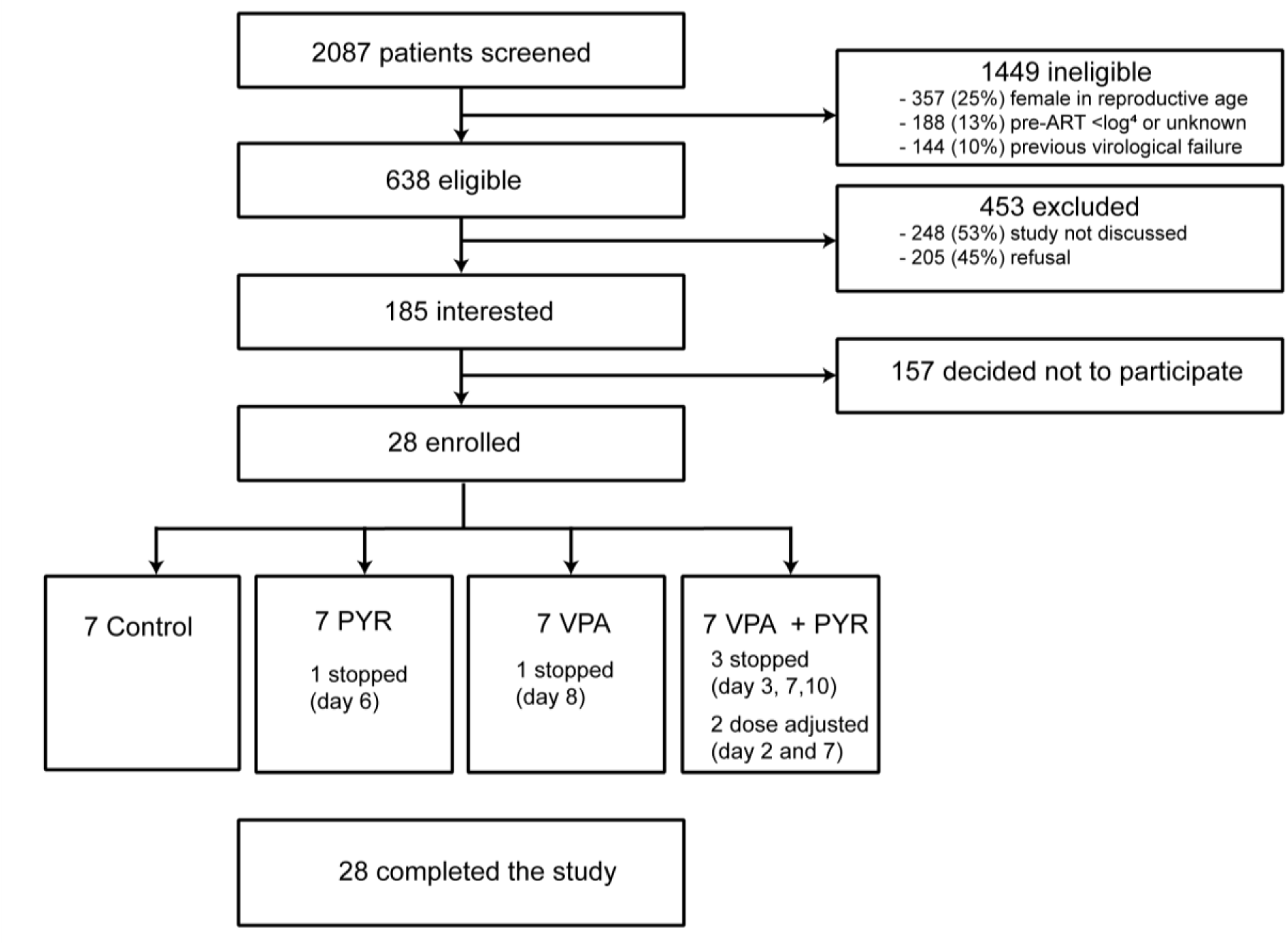
Study flow diagram showing the information about the method of recruitment and the number of participants that completed the LUNA study. Miscellaneous category for ineligibility covers not fitting the other study in- and exclusion criteria, clinical judgement of treating physician, and study not discussed. ART: antiretroviral therapy, PYR: pyrimethamine, VPA: valproic acid.

**Table 1.** Baseline characteristics of the participants at inclusion.

| **Baseline Characteristics** | **Overall**<br>**n =28** | **PYR**<br>**N=7** | **VPA**<br>**N=7** | **PYR+VPA**<br>**N=7** | **Control**<br>**N=7** |
| --- | --- | --- | --- | --- | --- |
| Male | 28 (100) | 7 (100) | 7 (100) | 7 (100) | 7 (100) |
| Ethnic origin |  |  |  |  |  |
| -White European | 24 (86) | 4 (43) | 7 (100) | 6 (86) | 7 (100) |
| -Latin American or<br>Hispanic | 3 (11) | 3 (57) | 0 | 0 | 0 |
| -Black Caribbean | 1 (4) | 0 | 0 | 1 (14) | 0 |
| Age (years) | 54 (47-61) | 53 (48-55) | 51 (54-67) | 51 (45-58) | 55 (51-64) |
| HIV subtype B<sup>a</sup> | 24 (85.7) | 7 (100) | 6 (86) | 6 (86) | 5 (71) |
| History of AIDS | 8 (29) | 2 (29) | 1 (14) | 3 (43) | 2 (29) |
| Years from HIV diagnosis<br>until inclusion | 8.5 (7.5-15.4) | 7.7 (7.5-14.5) | 7.8 (7.6-12.8) | 7.6 (5.2-9.4) | 15.6 (10.5-20.2) |
| Years on cART | 7.5 (5.6-11.7) | 7.3 (6.4-12.0) | 7.6 (6.1-10.8) | 5.6 (4.7-7.4) | 9.8 (8.0-13.0) |
| Years with HIV-RNA<br><50 copies per mL | 6.8 (4.9-11.1) | 6.7 (5.1-11.5) | 7.2 (5.4-9.5) | 5.1 (4.5-7.1) | 7.2 (5.2-12.5) |
| Initiated cART during<br>acute HIV infection | 3 (11) | 2 (29) | 1 (14) | 0 | 0 |
| Pre-cART plasma viral<br>load zenith log<sub>10</sub> copies<br>per mL | 4.9 (4.7-5.3) | 5.3 (4.9-5.6) | 4.8 (4.6-5.0) | 4.9 (4.8-5.0) | 4.9 (4.7-5.4) |
| Pre-cART nadir CD4+ T-<br>cell count per µL | 235 (160-315) | 290 (135-310) | 250 (225-295) | 240 (175-420) | 210 (170-225) |
| CD4+ T-cell count per µL<br>at inclusion | 665 (530-820) | 610 (490-770) | 750 (560-810) | 630 (475-1005) | 680 (590-740) |
| cART |  |  |  |  |  |
| -NNRTI based<sup>b</sup> | 16 (57) | 5 (71) | 4 (57) | 4 (57) | 3 (43) |
| -INSTI based<sup>c</sup> | 12 (43) | 2 (29) | 3 (43) | 3 (43) | 4 (57) |

Data are number with percentage or median with interquartile ranges.
<sup>a</sup>Other HIV-1 subtypes include CRF01-AE (n=2), CRF01-AG (n=1), C (n=1).
<sup>b</sup>NNRTI were rilpivirine (n=6), nevirapine (n=6), efavirenz (n=4). 15 NRTI backbones consisted of emtricitabine (FTC) and either tenofovir disoproxil fumarate (TDF) or tenofovir alafenamide (TAF) and 1 NRTI backbone consisted of abacavir with lamivudine.
<sup>c</sup>INSTI were dolutegravir (n=10) and elvitegravir/cobicistat (n=2). 7 NRTI backbones consisted of FTC and either TDF or TAF, 3 NRTI backbones consisted of lamivudine (3TC) and abacavir (ABC); and 2 had dolutegravir with 3TC as NRTI backbone.
Abbreviations: cART: combination antiretroviral therapy; INSTI: integrase strand transfer inhibitor; NNRTI: non-nucleoside reverse transcriptase inhibitor.

During the 6-week study duration after inclusion, a total of 103 adverse events (AE) were reported by the trial participants (**Table S1 and S2**). Except for three grade 3 AE (two with pyrimethamine alone, one in the combination arm), all were of grade 2 or lower severity (**Table S3**). Most AE (84/103) were experienced on pyrimethamine at a comparable rate between the monotherapy and the combination arms. Neurological and gastro-intestinal symptoms were the most common AE categories across all interventional arms with nausea, vomiting and headache being the most frequently reported. No serious adverse events (SAEs) were observed during the trial and all AE resolved after the intervention. No AIDS-defining illnesses occurred during or after the study. The blood CD4+T-cell counts remained stable during the study without CD4+T-cell counts below the predefined safety limit of 200/μl. However, reported grade 1 and 2 AE in seven of the participants necessitated the trial physicians to adjust the allocated treatment regimens. The PLWH with and without dose adjustments had a largely comparable clinical profile (**Table S4**). Three participants in the combination arm stopped treatment at day 3 (LUNA-23), day 7 (LUNA-06) and day 10 (LUNA-21), one participant in the pyrimethamine arm stopped treatment at day 6 (LUNA-10) and one participant in the valproic acid arm stopped treatment at day 8 (LUNA-09). In one participant of the combination arm we halved pyrimethamine dosage at day 7 (LUNA-12), while in another participant in the combination treatment arm we halved the valproic acid dose at day 2 and pyrimethamine dose at day 7 (LUNA-25) due to drug related AE. These events met the upper limit of our predefined safety criteria where we would stop the trial if more than two PLWH in a treatment arm had to discontinue treatment when 50% inclusion was reached, or if at any moment in the trial more than five PLWH had to discontinue their treatment for drug related AE. We concluded that there was sufficient clinical-scientific and medical ethical basis to complete the study and had 100% follow up of all 28 participants.

### Pyrimethamine induces HIV-1 transcription *in vivo*

To assess the potential of HIV-1 latency reversal in PLWH induced by pyrimethamine and valproic acid alone and in combination, we measured cell associated unspliced (CA US) HIV-1 RNA by RT-qPCR in CD4+T-cells isolated from peripheral blood at baseline (day 0, t=0 hours), 6 hours after initiating the intervention regimen (day 0, t=6 hours), at the end of the 2-week intervention phase (day 14), and 4 weeks thereafter (day 42) (**Fig. S1**). Due to low cell numbers in the baseline sample, LUNA-06 (combination arm, participant stopped at day 7) was excluded from this primary endpoint analysis.

Addressing the primary endpoint of the study, we found that, overall, the absolute change in CA US HIV-1 RNA from baseline during the 6-week study period between the treatment groups was different (P<0.001) (**Fig. 2A**). This overall analysis cannot be used to conclude which groups or timepoints determined the effect, but the graphical data suggested a relevant contribution of the groups exposed to pyrimethamine. We performed additional analyses to substantiate this possibility. Overall, the fold change in CA US HIV-1 RNA is significantly induced in the pyrimethamine arm at day 14 compared to the control arm (**Fig. 2B, Table 2, Table S5**). Induction of CA US HIV-1 RNA in the pyrimethamine arm was consistently more prominent compared to controls, although not reaching statistical significance at all timepoints, likely due to the sample size (**Table 2**).

**Fig. 2.**
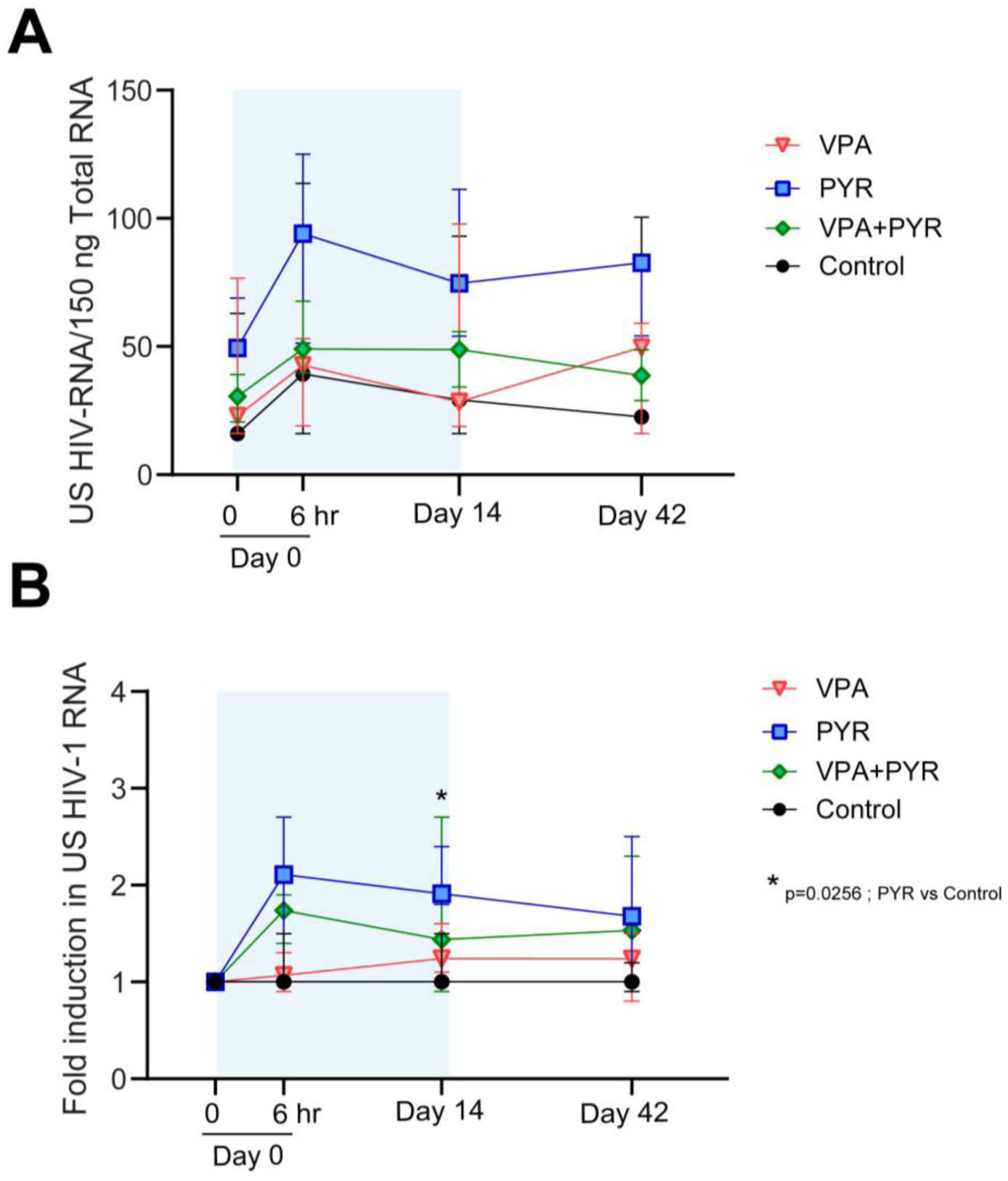
Changes in CA HIV-1 US RNA per arm at four time points during the LUNA study. (A) median (interquartile range) total CA US HIV-1 RNA per 150 ng of total RNA in all four study arms at treatment initiation (day 0, t=0hr) and 6 hours after first dosing (day 0 t=6hr), at the end of the treatment period (day 14) and 28 days after the end of treatment (day 42). (B) Median (interquartile range) fold change in CA US HIV-1 RNA in all four study arms relative to baseline (day 0, t=0hr). * indicates the effect comparisons at the timepoints of the treatment arms compared to controls with p<0.05.

**Table 2.**
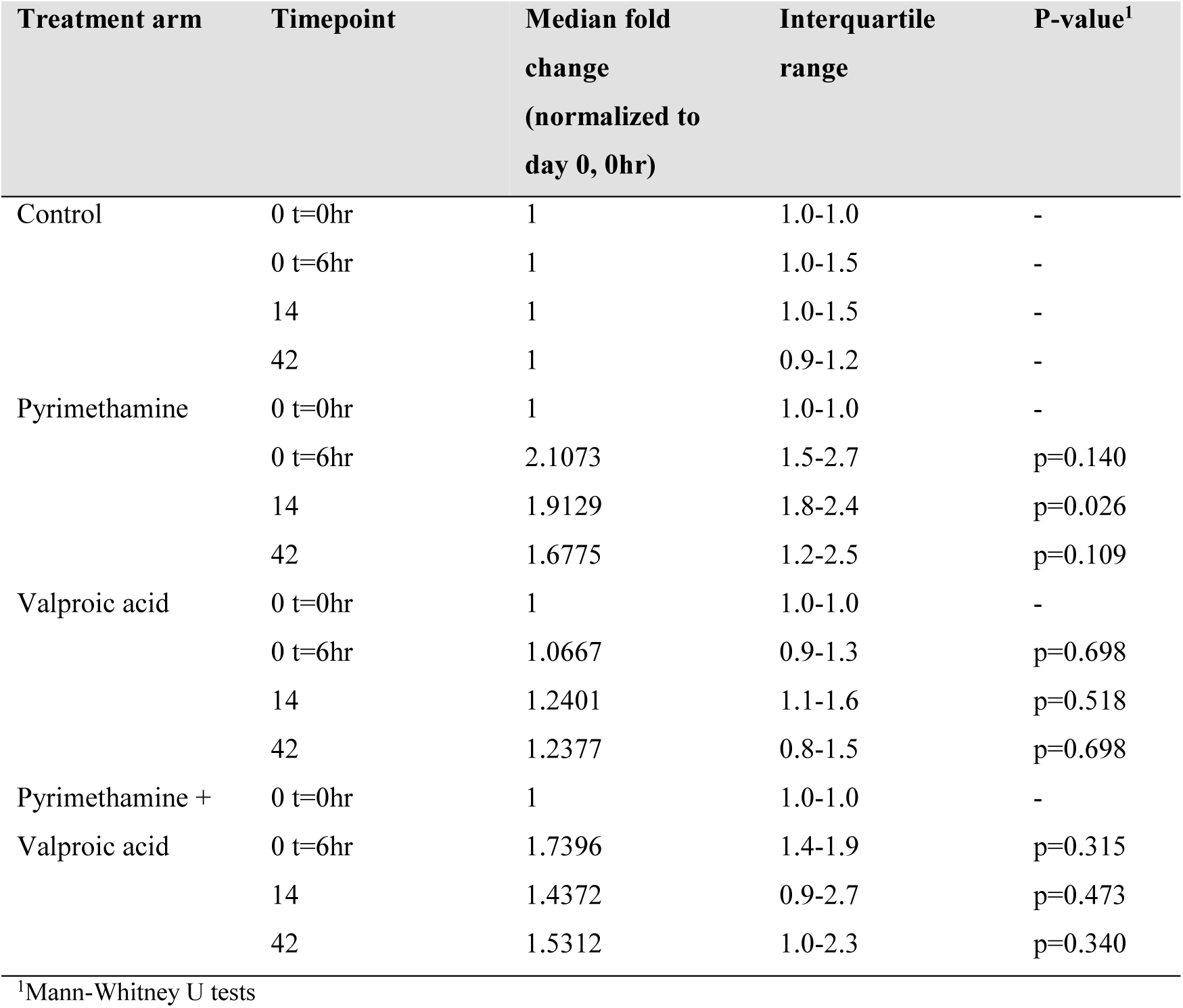
The median fold change in CA US HIV-1 RNA per timepoint and per treatment arm compared to the control arm.

As a consequence of the treatment adjustments, the 6 hours timepoint after the first dosing is the only sampling timepoint where all patients were similarly exposed to their allocated therapies and at the same doses. We therefore focused on the effects observed here as the most unbiased measurement timepoint consequent to drug treatment. From baseline to 6 hours, CA US HIV-1 RNA increased median 2.1 fold (IQR: 1.5-2.7, p=0.028) in response to pyrimethamine alone (**Fig.3A and 3E**) and median 1.7 (IQR: 1.4-1.9, p=0.046) in response to the combination therapy (**Fig. 3B and 3F).** In contrast, the CA US HIV-1 RNA at this timepoint from baseline did not change significantly for PLWH allocated to the valproic acid arm (median fold change 1.1 IQR: 0.9-1.3, p=0.46) (**Fig. 3C and 3G**) or the control arm (median fold change 1.0, IQR: 1.0-1.5, p=0.14; **Fig. 3D and 3H**). Overall, PLWH in the two arms exposed to pyrimethamine had a greater fold change CA US HIV-1 RNA after 6 hours (median 1.8, IQR: 1.4-2.4) compared to those in the two arms without pyrimethamine exposure (median 1.0, IQR: 1.0-1.4; p=0.029).

**Fig. 3.**
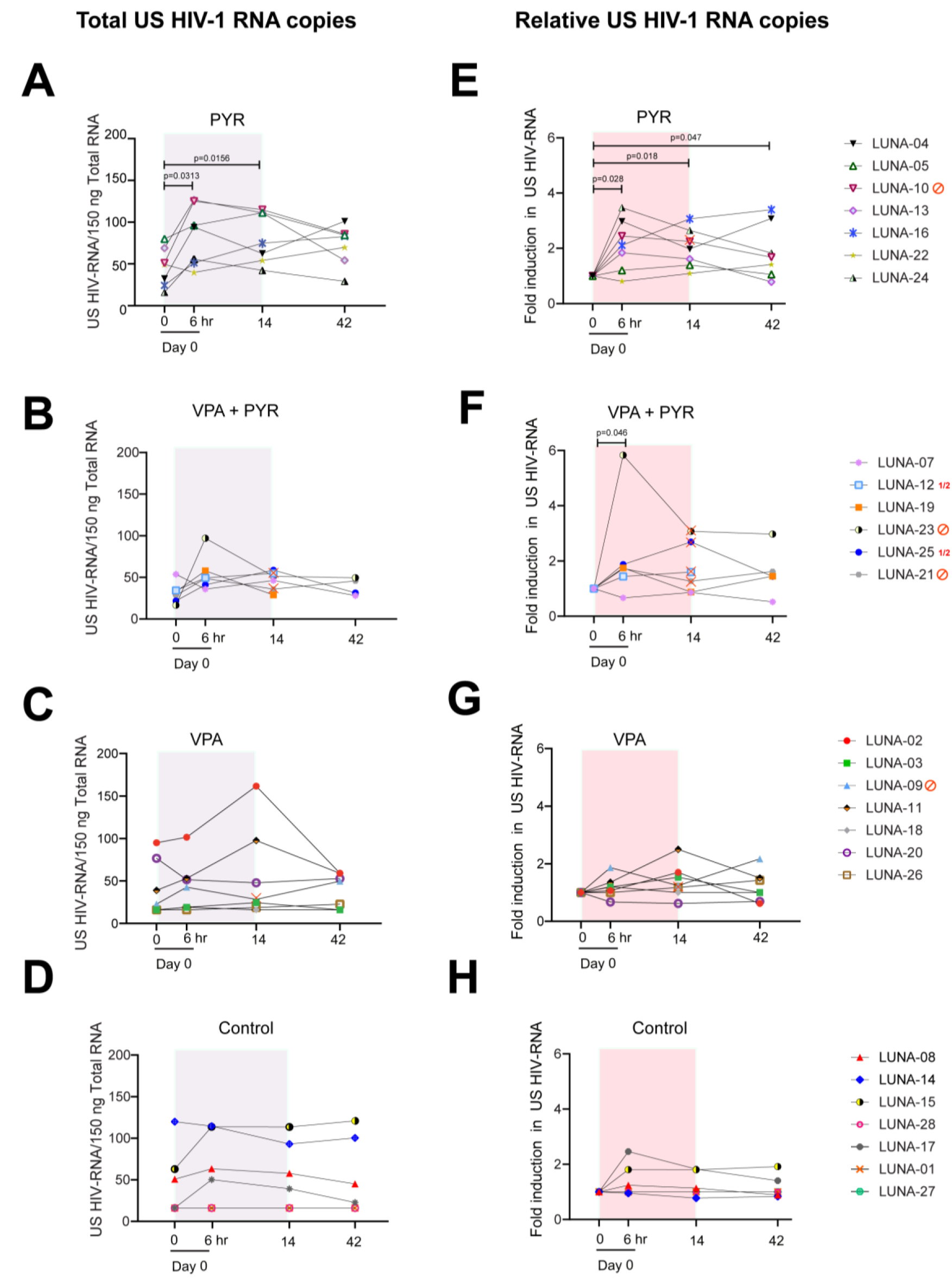
Changes in CA HIV-1 US RNA per participant per arm at four time points during the LUNA study. (A-D) Graphs represent absolute CA US HIV-1 RNA copies in all participants from the pyrimethamine (A), pyrimethamine with valproic acid (B), valproic acid (C) and control (D) arms at treatment initiation (day 0, t=0hr) after 6 hours after first dosing (day 0, t=6hr), at the end of treatment period (day 14) and 28 days after end of treatment (day 42). (E-H) Graphs represent fold induction of CA US HIV-1 RNA at different study time points compared to baseline (day 0, t=0) in all participants from the pyrimethamine (E), pyrimethamine with valproic acid (F), valproic acid (G) and control (H) arms at the same timepoints. 1.5 million CD4+ T cells were isolated from PBMCs, lysed in triplicate, followed by total RNA isolation and reverse transcription. Total copies of CA US HIV-1 RNA were quantified by nested qPCR. The grey/red boxes represent treatment duration. Participants that stopped study medication or had dose adjustments are indicated with either a red stop sign or red ½ symbol and a red cross on the individual points in the graph. In both the valproic acid arm and the pyrimethamine arm, one participant stopped their study medication on day 8 (LUNA-09) and day 6 (LUNA-10), respectively. In the combination treatment arm, three participants stopped both compounds (LUNA-23 on day 3, LUNA-06 on day 7 and LUNA-21 on day 10), and two participants had their dosage adjusted (LUNA-25 had VPA dose halved from day 2 on, PYR dose halved from day 7 on, and LUNA-12 had PYR dose halved from day 7 on). P-values below 0.05 are indicated. Statistical significance was calculated using Wilcoxon Signed Rank Tests.

On pyrimethamine monotherapy, six out of seven PLWH responded after the first dose at the 6 hour timepoint (**Fig. 3A and 3E**). Of these six, five retained an increased CA US HIV-1 RNA during pyrimethamine treatment including LUNA-24 who had CA-US HIV-1 RNA below the level of quantitation at baseline. The median fold change from baseline at day 14 was 1.9 (IQR: 1.5-2.4, p=0.018), which was in line with the median fold change at 6 hours after dosing (p=0.612) and remained comparable (p=0.50) from day 14 to day 42 when a median 1.7 fold change (IQR:1.2-2.5) was still observed. Excluding the single patient (LUNA-10) who stopped pyrimethamine from these analyses did not relevantly alter these median fold change estimates from baseline at day 14 (1.8 IQR:1.4-2.6) or day 42 (1.6, IQR: 1.1-3.1). Controls did not exhibit relevant median fold change increases in CA US HIV-1 RNA from baseline at day 14 (1.0, IQR: 1.0-1.5, p=0.27) or day 42 (1.0, IQR:0.9-1.2, p=0.47). Two PLWH in the control arm showed some reactivation without a clear clinical explanation, which skewed the estimated effect ranges upwards. On valproic acid, PLWH did not have significantly increased median CA US HIV-1 RNA at any timepoint compared to pretreatment levels (**Fig. 3C and 3G**) and these responses were also similar to the responses measured in untreated controls (p>0.5 for all comparisons). Together these observations strongly indicate that pyrimethamine induces HIV-1 transcription as early as 6 hours after intake.

We did not observe plasma viremia despite the observed increases in CA US HIV-1 RNA. We found no evidence that pyrimethamine induced plasma viremia at the day 1 timepoint when all PLWH were on their allocated treatments; five PLWH exposed to pyrimethamine and four PLWH not exposed to pyrimethamine had plasma HIV-1 RNA detected below the assay quantitation limit. These included six PLWH, equally divided by pyrimethamine exposure or not, who had no viral genome detected prior to drug administration. When evaluating the entire 6 week trial period, plasma HIV-1 RNA remained below the lower limit of quantification in the majority of participants (**Table S6**) except for two participants on valproic acid, one participant on pyrimethamine and one participant on combination treatment (all <50 copies/mL). These findings indicate that the intracellular effect of pyrimethamine did not result in measurable cell free HIV-1 RNA in the plasma of participants.

### Pyrimethamine and valproic acid do not synergize to induce HIV-1 transcription *in vivo*

Due to the treatment adjustments that occurred in the combination treatment arm, we used the measurements at 6 hours after the first dose to test the hypothesis that synergy occurred between pyrimethamine and valproic acid in the context of their combined latency reversal activity on HIV-1. In PLWH exposed to the combinatorial treatment, five out of six PLWH had increased CA US HIV-1 RNA 6 hours after the first dose (**Fig. 3B and 3F**). There was one individual (LUNA-23) with a particularly large 5.83 fold increase while the other five PLWH had fold changes ranging 0.67 to 1.88. Apart from an HIV-1 CRF02-AG subtype infection, the clinical characteristics of this patient were fitting the characteristics of the overall study population. At this 6 hour timepoint, the median fold change response in the combination treatment arm did not differ significantly from the median response observed with pyrimethamine alone (p=0.475). As per protocol defined, the data also formally excluded synergy since the mean fold change and its full 95% confidence interval (2.2, 0.3-4.1) did not exceed the expected mean fold change of 2.25, which was based on the mean fold change effects estimated in the pyrimethamine (2.11) and valproic acid (1.14) arms separately. Hence, synergy could not be demonstrated and neither do these data indicate additive effects.

In the four PLWH (LUNA-07, LUNA-12, LUNA-19, LUNA-25) in the combination arm who continued uninterrupted pyrimethamine exposure throughout the study, the median fold change in CA US HIV-1 RNA was 1.2 (IQR: 0.9-2.1) at day 14. This effect was comparable to the changes observed when evaluating all PLWH including those who stopped combinatorial treatment (median 1.4, IQR: 0.9-2.7). Unfortunately, the CD4+T-cell yield from the samples were insufficient for LUNA-12 and LUNA-19 to measure CA US HIV-1 RNA at the day 42 endpoint. Given the treatment adjustment in LUNA-25, at this timepoint only LUNA-07 had completed the full allocated treatment and had endpoint results available. This patient did not exhibit CA US HIV-1 RNA increases at any timepoint. Apart from a longer duration of plasma viral suppression in the upper quartile of the cohort (nearly 12 years), the other baseline characteristics in this participant were consistent with the general cohort. When evaluating the four PLWH who had exposure to pyrimethamine in this combination treatment arm and with samples at day 42 available (LUNA-07, LUNA-21, LUNA-23, LUNA-25), the median fold change was 1.5 (IQR: 1.0-2.3), comparable to pyrimethamine alone. Overall, these data support the conclusion that valproic did not augment the activity of pyrimethamine in reactivating HIV-1.

### Pharmacokinetics of pyrimethamine and valproic acid

For this trial, we established an in-house assay to measure total pyrimethamine plasma levels (**Fig. 4**). At the 6 hour timepoint, the median plasma total pyrimethamine concentration in the seven participants that received solely pyrimethamine was 1.63 mg/L (IQR 1.40-1.83) and in the combination arm this was 1.16 mg/L (IQR: 1.0-1.31) (**Fig. 4A**). This difference was more accentuated at day 14 when the six PLWH in the monotherapy arm and the four PLWH in the combination therapy arm with uninterrupted pyrimethamine exposure had median C_trough_ of 3.48 mg/L (IQR: 3.21-3.83) and 1.99 mg/L (1.63-2.63) (**Fig. 4B**). Although these concentrations fall within the expected range from previous human studies (*49–52, 58*), and despite taking out those who stopped pyrimethamine before day 14 from the analysis, the data suggest lower pyrimethamine concentrations when combined with valproic acid (**Fig. 4**). This might be the result of protein binding displacement or an unexpected drug-drug interaction. We did not find such an effect when comparing the median plasma total valproic acid concentration at the 6 hour timepoint between monotherapy (median 47.3 mg/L, IQR: 47.2-60.1) and combination treatment (47.4 mg/L, IQR: 26.0-57.3) (**Fig. S2**). The median C_trough_ observed at the day 14 timepoint in the six PLWH with uninterrupted exposure to valproic acid monotherapy and the four PLWH with uninterrupted exposure to valproic acid in the combination treatment were 84.8 mg/L (36.4-99.6) and 113.2 mg/L (IQR: 83.1-137.5) respectively, both above valproic acid’s therapeutic range lower border for epilepsy (50 mg/L). We found no correlations between pyrimethamine plasma exposure and the fold change in CA US HIV-1 RNA from baseline at 6 hours (r=0.26, p=0.39) or day 14 (r=0.041, p=0.91) in those exposed to pyrimethamine or in the participants from the two pyrimethamine arms separately (**Fig. S3**). We explored median pyrimethamine C_trough_ by integrase inhibitor exposure but found no relevant differences at 6 hours (1.3 mg/L vs 1.4 mg/L) or day 14 (2.8 mg/L vs 3.2 mg/L) in those with uninterrupted pyrimethamine. Conversely, median unbound dolutegravir concentrations were comparable between those with and without pyrimethamine exposure at 6 hours (11.7 µg/L vs 10.1 µg/L), but decreased during the treatment course. This effect has been attributed to valproic acid by protein displacement as we described previously (*59*). PLWH with exposure to efavirenz or nevirapine-containing cART (both CYP3A inducers) did not have lower median plasma concentration of pyrimethamine (a CYP3A substrate). No pyrimethamine or valproic acid was detectable in the plasma at day 42. The combined data do not support relevant drug-drug interactions between pyrimethamine and dolutegravir but do support relevant effects by valproic acid on dolutegravir and pyrimethamine.

**Fig. 4.**
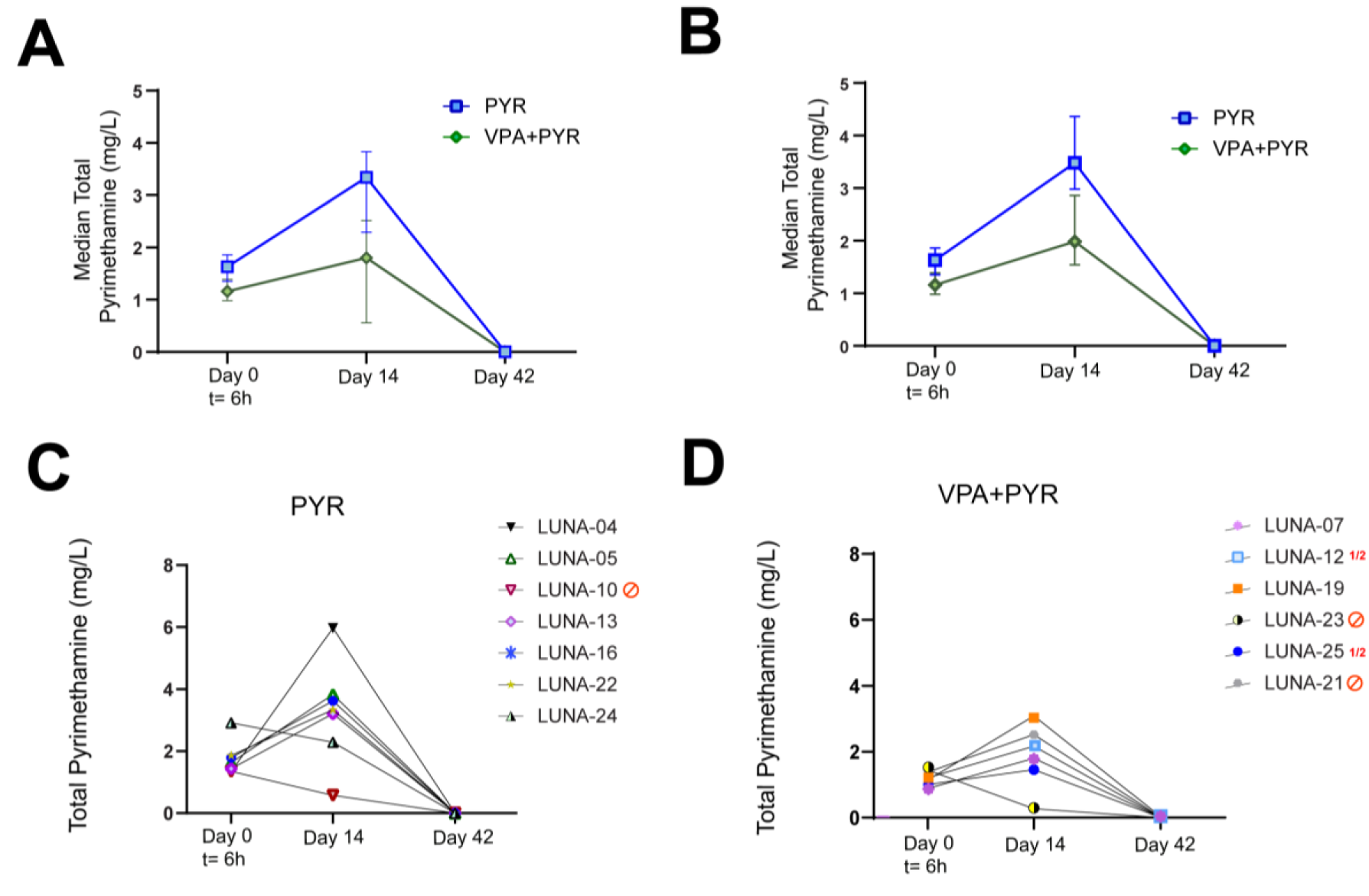
Pharmacokinetics of the drug pyrimethamine in study participants. (A-B) Pharmacokinetics analysis of the drug pyrimethamine in study participants from pyrimethamine (blue) and the combination treatment arm with pyrimethamine and valproic acid (green) as measured by median total pyrimethamine (mg/L) levels in plasma with interquartile ranges at 6 hours after first dosing (day 0, t=6hr), at the end of treatment period (day 14) and 28 days after end of treatment (day 42) overall (A) and in those with uninterrupted pyrimethamine exposure (B) and represented as plasma pyrimethamine levels at the timepoints per individual on pyrimethamine (C) or the combination treatment with pyrimethamine and valproic acid (D).

We found no clear outliers in the data on pyrimethamine and valproic acid plasma exposure for the seven PLWH who had treatment adjustments for toxic effects (**Table S6**). Of those seven, the ones in the pyrimethamine containing arms had pyrimethamine plasma levels at 6 hours after the first dose that were at or below the median plasma exposure found in the pyrimethamine monotherapy arm at that timepoints. All patients where pyrimethamine was stopped had clearly lower plasma levels at day 14 with the notable exception of LUNA21 (2.52 mg/L) who stopped treatment at day10, later than the others who interrupted pyrimethamine (**Table S6**). In the same line, three of the five PLWH, who had their therapy adjusted for toxicity while receiving valproic acid, had plasma levels at day 7 exceeding the therapeutic window defined for treating epilepsy (up to 100 mg/L) compared to three out of nine who continued valproic acid (**Table S6**). This provides some evidence that valproic acid plasma exposure related to clinical toxicity, comparable to the use of therapeutic drug monitoring of valproic acid for other conditions (*60*). For pyrimethamine, the therapeutic window and relationship between plasma exposure and toxicity are far less well defined but no exceptional high plasma exposures were found.

### Pyrimethamine induces expression of biomarkers associated with BAF inhibition

Pharmacological inhibition of the BAF complex by pyrimethamine leads to functional changes in the expression levels of several genes (*44, 61–64*). To assess the pharmacodynamics, activity and specificity of pyrimethamine in this clinical trial, we analyzed the gene expression profile of three target genes of the BAF complex (IL-10, SOCS3, CBX7) and a control gene (B-ACT) on day 0 before treatment and day 0 at 6 hours after the first dose. We reasoned that the effect on the expression of these gene targets at later timepoints (day 14 and day 42) would be less useful as specific biomarker of BAF complex inhibition due to treatment interruptions and potential induction via other pathways. We observed a significant fold increase in the mean gene expression levels of IL-10, SOCS3 and CBX7 (3.14, 2.82 and 3.39-fold respectively) in the pyrimethamine treatment arm (**Fig. 5A**). As expected, we observed no increase in the mRNA levels of these genes in neither the control arm nor the valproic acid intervention arm (**Fig. 5B-C**), indicating that these molecular targets are specific for pyrimethamine. Interestingly, only a moderate increase in gene expression levels of BAF target genes were observed in the combined intervention arm (**Fig. 5D**). These findings are consistent with, and might be the consequence of, the generally lower plasma pyrimethamine C_trough_ in participants that received the combined intervention regimen compared to those that received solely pyrimethamine.

**Fig. 5.**
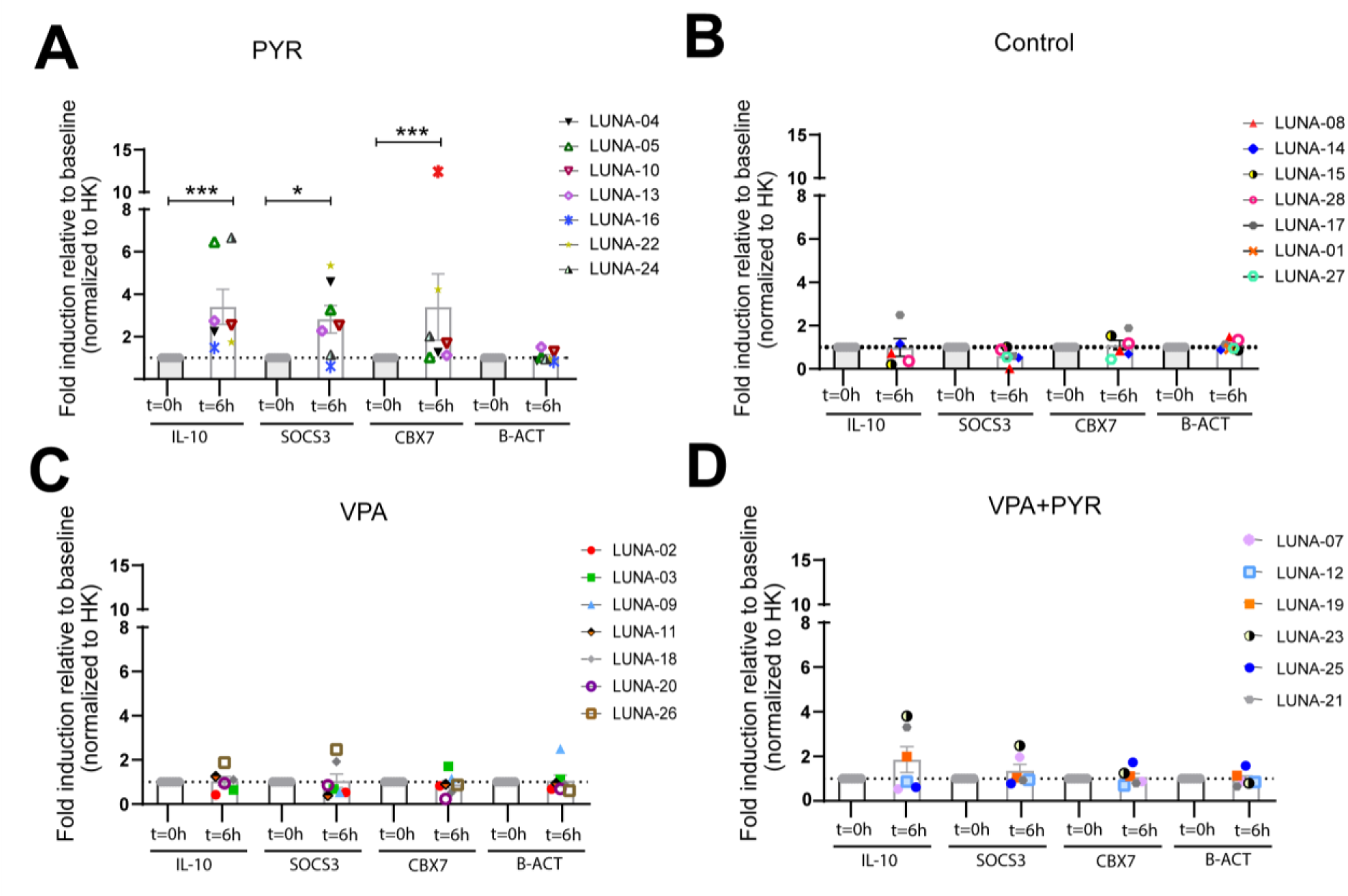
Pharmacodynamics of the drug pyrimethamine in study participants. (A-D) Gene expression profile of BAF complex molecular targets IL-10, SOCS3, and CBX7 in all participants from pyrimethamine (PYR, A) control (B), valproic acid (VPA, C) and the combination treatment arm with pyrimethamine and valproic acid (D) at 6 hours after first dosing (day 0, t=6hr). Graphs represent mean fold induction relative to pretreatment level (day 0, t=0hr). Cyclophilin A was used as a housekeeping gene (HK) for normalization and beta-actin (B-ACT) was used as a control. P=values were calculated using unpaired t-test with *representing p<0.05 and ** representing p<0.01.

### Pyrimethamine exposure does not lead to a reduction of the inducible HIV-1 reservoir

To assess whether *in vivo* HIV-1 transcription induced by pyrimethamine or valproic acid alone or in combination impacted the inducible viral reservoir, we used a tat/rev induced limiting dilution assay (TILDA) to quantify the frequency of cells expressing multiply spliced (MS) HIV-1 RNA upon ex vivo cellular activation with PMA/Ionomycin at baseline (day 0, t=0hr) and day 42 (**Fig. 6 and Supp. Fig. S4A-C**) (*65, 66*). Since low cell yields at day 42 occurred in several individuals during the trial, which would impede accurate reservoir assessments, we decided during the trial, before data collection and analysis, to invite all participants back when all were more than 1 year after day 42. A total of 27/28 PLWH responded and were sampled at a 1 – 2.2 years range after their last visit (**Table S7**). Ultimately, TILDA could be performed for 20/28 study participants with available samples (71%) including all participants on pyrimethamine monotherapy. For eight participants without TILDA data, sample material was insufficient in four (three on combination treatment and one control) and four had non-B HIV-1 subtypes (including one participant in the combination arm). For the remaining ten participants with pyrimethamine exposure that could be evaluated, two on pyrimethamine alone (LUNA-05 and LUNA-22) had insufficient cells available at day 42. Here, the measurement from the cells obtained >1 year after the last visit were used.

**Fig.6.**
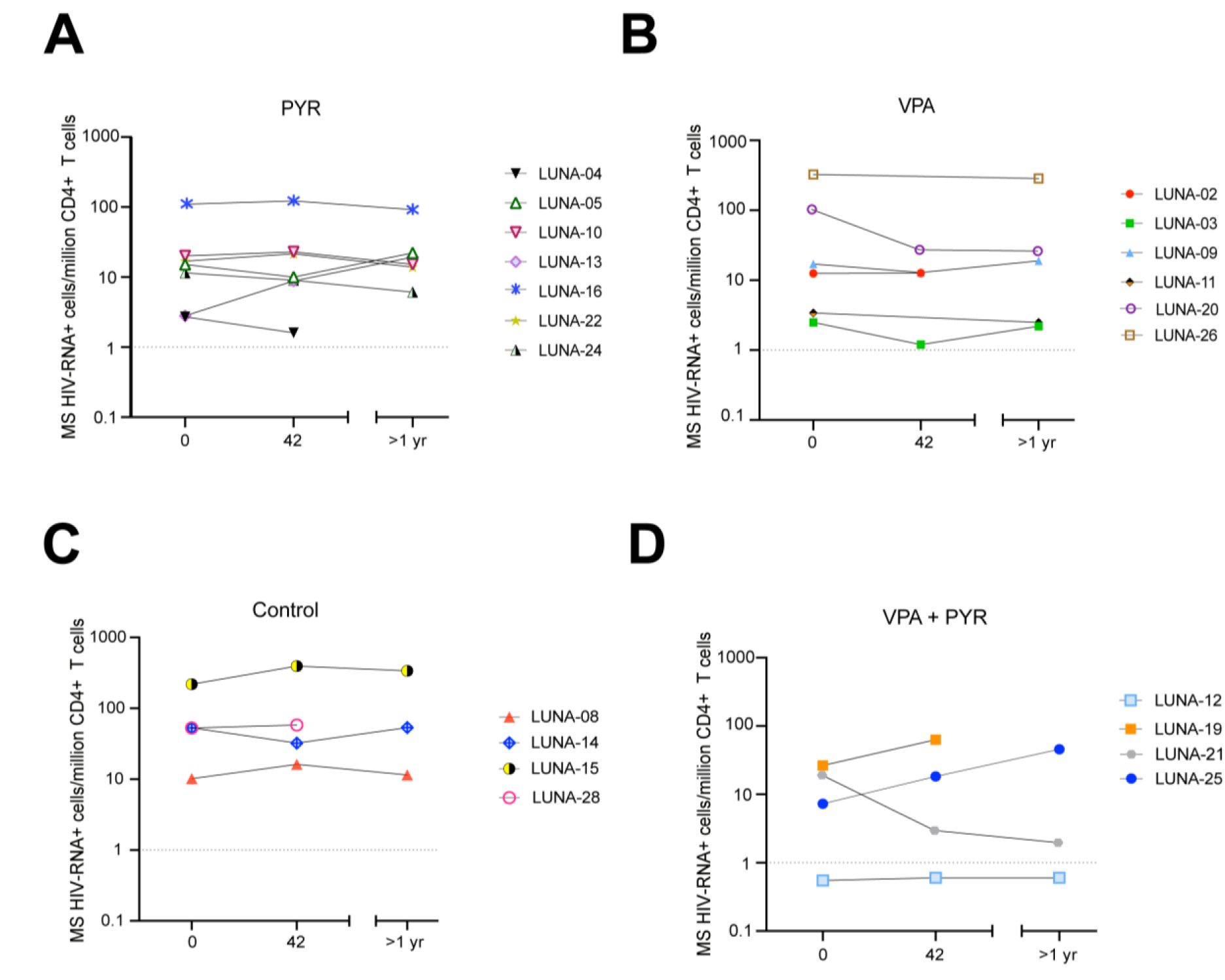
Effect of the intervention regimen on the HIV-1 inducible reservoir size. (a-d) Graphs represent measurements of inducible reservoir size in samples obtained from participants from pyrimethamine (A), valproic acid (B), control (C) and the combination treatment arm with pyrimethamine and valproic acid (D) at treatment initiation (day 0, t=0hr), 28 days after end of treatment (day 42) and >1 year after the end of treatment. Isolated CD4+ T cells were activated ex vivo with PMA/ionomycin for 12 hours and the frequency of CD4+ T cells expressing multiply spliced (MS) HIV-1 RNA was determined using a *tat/rev* induced limiting dilution assay (TILDA). The dotted horizontal line represents the assay limit of detection.

The overall results do not indicate a significant change in the reservoir size with pyrimethamine exposure (**Fig. 6A**). The reservoir was estimated at median 15.1 (range 2.7 to 109.8) cells/million CD4+T-cells before treatment with pyrimethamine alone and at 10.0 (range 1.6 to 122.8) cells/million CD4+T-cells after treatment. This variation was well within the assay coefficient of variation (25.1%). The reservoir remained stable on valproic acid; median 14.8 cells/million CD4+T-cells at day 0 and median 19.4 CD4+T-cells /million at day 42 (**Fig. 6B**). The number of participants with reservoir results available in the other two arms (n=3 in each) were considerably lower and precluded statistical inferences but the data points that are available do not challenge the main conclusion that overall no decrease in the reservoir can be observed (**Fig. 6C-D**). In one of these participants (LUNA-12, combination arm), TILDA was below the limit of detection at all sampling points (**Fig. 6D**).

## DISCUSSION

The BAF complex is an important molecular regulator of HIV-1 latency and offers a new yet clinically unstudied pharmacological target for latency reversal. In this randomized controlled trial, we demonstrated that BAF complex inhibition by pyrimethamine induces cellular HIV-1 transcription and reversal of latency in cART-suppressed PLWH. A comparable effect on the reservoir by pyrimethamine was also observed when valproic acid was added as partner LRA. Thus, the combination treatment failed to show synergistic or additive effects. Furthermore, interventional treatment adjustments occurred most frequently in PLWH randomized to the combinatorial arm, although the number and grade of the reported AE were in line with the ones observed in the individual monotherapy arms. We cannot exclude the possibility of physician or patient-directed biases in the combinatorial arm where both the treating physician and the participant were aware of the higher drug burden. However, we believe that this trial, as the first randomized controlled clinical study on combinations of LRA, underlines that testing combinatorial LRA approaches is feasible. It also highlights the need to monitor for drug tolerability and provides a safety benchmark for future studies combining LRAs in the clinical HIV-1 cure trial field.

The most important finding is the identification of pyrimethamine as an effective drug to reverse HIV-1 latency in PLWH. It acts through a novel mechanism of action that we unraveled first in *in vitro* and *ex vivo* studies and now successfully moved into clinical practice (*44, 48*). We found an approximately 2-fold induction of cellular HIV-1 transcription from pre-treatment levels that persisted during treatment. The effect on the proviral reservoir was only observed in the presence of measurable pyrimethamine plasma concentrations, accompanied by selective induction of other BAF complex target genes and not observed in individuals that were not exposed to pyrimethamine. These observations support that the inhibition of the BAF complex at the HIV-1 promotor region is responsible for this effect *in vivo*, comparable to its established working mechanism *in vitro*. In addition, although BAF complex inhibition by pyrimethamine was able to induce HIV-1 transcription in vivo, it did not lead to a reduction in the inducible reservoir. This might be due to an insufficient antigen production and further recognition by cytotoxic immune cells.

With regard to valproic acid, we optimized the dosing and used an acid resistant formulation for a more direct gastrointestinal absorption unlike the previous studies where under-dosing or -exposure might have been the cause of the observed discrepant effects of valproic acid as LRA (*17–22*). In our trial, participants receiving valproic acid had higher plasma levels than those reported in prior studies (*17–22*). Although higher systemic valproic acid peak concentrations could more likely induce HIV-1 reactivation, we did not observe an increase in CA US HIV-1 RNA or viral plasma load in PLWH treated with valproic acid alone.

Strategies that use combinatorial approaches of LRAs could result in higher efficacy in reactivating HIV-1 latency (*5*). We have shown in previous studies that the effect of pyrimethamine on HIV-1 latency *ex vivo* could be potentiated when combined with other LRAs including HDACi (*44, 46*). We therefore designed the study to include a combinatorial arm to test whether the effect of the individual LRAs could be potentiated *in vivo*. Our results showed, however, no synergistic or additive effects on the HIV-1 reservoir in PLWH in the combinatorial arm. Future HIV-1 cure studies using pyrimethamine would benefit from combinations with other LRAs that could induce higher levels of HIV-1 transcription and compounds that enhance immune-mediated killing.

To put our clinical study and the role of pyrimethamine as an LRA in context of HIV-1 cure clinical studies, we compared the effect of pyramethamine on HIV-1 latency with other LRAs tested in PLWH. Firstly, the effect of pyrimethamine on HIV-1 transcription did not appear to augment with multiple dosing. This is in contrast to observations made in studies with other LRAs from the HDACi class and disulfiram, where the mean effect on cellular HIV-1 transcription increased with sequential dosing during treatment (*26, 34, 39, 40*). Our observation fits a more dichotomous (on/off) effect dependent on whether sufficient intracellular concentrations are reached rather than a gradual effect seen with HDACi, where multiple doses are apparently necessary to reach a maximum effect. Our prior *in vitro* observations showing that the displacement of the BAF complex at the nucleosome 1 region of the HIV-1 promotor locus was comparable at different dosages supports this assumption (*44*). A certain maximized potential to reactivate HIV-1 is also suggested since a further increase in pyrimethamine plasma levels did not result in an additional CA US HIV-1 RNA increase during the trial. Secondly, pyrimethamine’s effect waned several weeks after last drug exposure. This is in line with most other trials on effective LRAs that act through de-repressing HIV-1 transcription (*34, 35, 39*) with the exception of one trial on vorinostat (*26*). Thirdly, the combinatorial approach used in this trial with pyrimethamine and a de-repressor HDACi as a partner LRA may be less likely to work synergistically compared to combining a de-repressing and an activating LRA. Our data support this hypothesis although we certainly acknowledge that different results may have been obtained with a more potent partnering HDACi. Future studies including pyrimethamine as an LRA should focus on optimizing the minimum necessary dose to reactivate HIV-1 latency while minimizing drug exposure and toxicity, and determine the optimal partner drug for pyrimethamine.

In our study pyrimethamine alone led to a 2-fold increase in levels of CA US HIV-1 RNA at the timepoint where all participants had received their allocated dose. While this effect is rather modest, it is comparable to recent reports using other classes of LRAs. The HDACi valproic acid did not result in an increase in CA US HIV-1 RNA in this trial, as opposed to what has been observed with other HDACi such as vorinostat, romidepsin and panobinostat (*26, 28, 33-39*). In a study more comparably designed to our study, CA US HIV-1 RNA increased a mean 2.6 fold at timepoints during vorinostat (*26*). No reactivation was however observed when vorinostat was combined with a T-cell inducing HIV-1 vaccine in PLWH who initiated cART during acute HIV-1 (*28*), and a 1.5 fold-change in CA US HIV-1 RNA was observed with vorinostat treatment in a recent study in postmenopausal women (*33*). For the HDACi romidepsin, initial studies reported an approximately 3-fold increase in CA US HIV-1 RNA (*35, 36*). Additional studies on romidepsin’s pharmacodynamics profile unexpectedly found no significant effect on HIV-1 transcription (*38*). In trials that combined romidepsin with 3BNC117 or a therapeutic HIV-1 vaccine, cellular HIV-1 transcript levels generally changed overall less than 3-fold (*37, 39*). This was also true for the reactivation effects observed with disulfiram, bryostatin, a TLR9 agonist or pembrolizumab (*40–43*). Another HDACi, panobinostat, reactivated cellular HIV-1 transcription more potent compared to all other HDACis in the only clinical trial with this compound (*34*). Overall, when comparing pyrimethamine to other LRAs, its effect to reactivate HIV-1 transcription can be classified as at least comparable.

The assay we developed and validated to measure pyrimethamine plasma levels turned out to be critical since unexpectedly, the combination arm had lower pyrimethamine plasma levels with a concomitant lower induction of various BAF target genes. Given that both pyrimethamine and valproic acid are highly protein-bound, a plausible explanation for this effect could be attributed to plasma protein-protein competition dynamics, although further research needs to be conducted to support this statement. We identified a similar mechanism previously where valproic acid affected dolutegravir levels (*59*). This signals that routine pharmacokinetic profiling of the interventional drugs and cART should be included in the design of future cure trials, even if no interaction is expected. This is particularly important when testing combinations of LRAs.

This proof-of-concept study has strengths and limitations. The study’s main strength is the inclusion of a control arm and a combination arm that allowed us to assess the specific effect of pyrimethamine alone and in combination with an HDACi. Regarding limitations, although not exceeding predefined safety criteria, the combinatorial arm faced a disproportionate amount of treatment adjustments before the end of the intervention period. This influenced our ability to draw solid conclusions on potential late LRA effects combination therapy might have. However, the absence of an effect with valproic acid alone together with the absent synergism or additive effects in PLWH on the combination of drugs make potential late effects less likely. Our sampling strategy for this pilot favored having many timepoints for the primary endpoint over blood quantity for in-depth analyses. Leukapheresis was practically not possible. This turned out to be a challenging strategy in some cases, especially in individuals with less blood CD4+T-cells. We prioritized using CD4+T-cells for the main endpoints although this meant that in depth analysis of resting memory CD4+T-cells or other cell types was therefore not possible. Another challenge was to measure the size of the reservoir by TILDA post-treatment at day 42 as a main secondary endpoint. This was not possible in four PLWH due to limited sample availability and four PLWH had non-B subtypes that resulted in critical primer-probe mismatches. However, all PLWH in the most relevant arm with pyrimethamine had a sufficient amount of cells for reservoir analysis. We amended the protocol and invited all participants for an additional sampling at least 1 year after the last study visit with the purpose to more confidently support the reservoir findings. We did not observe marked changes in the size of the latent reservoir over this timecourse. Future LRA trial should take into account these limitations for initial design with regards to diversity in participant inclusion, assays, sampling timepoints and materials required.

In summary, inhibition of the BAF complex by pyrimethamine resulted in modest but significant reversal of HIV-1 latency in PLWH immediately after the first dose and persisted during treatment course. Our data is supportive of the BAF inhibitor Pyrimethamine as a novel drug option in the LRA arsenal, which has widespread use in the clinic, excellent safety profile and favorable pharmacological features, including its excellent brain penetration that offers potential for penetration in HIV-1 reservoir sanctuary sites (*50, 53, 54, 56, 58*). Thus Pyrimethamine is an attractive candidate for inclusion in future pharmacological approaches towards an HIV-1 cure.

## MATERIALS AND METHODS

### Eligibility

The LUNA trial (clinicaltrials.gov identifier: NCT03525730) is a proof of concept, four arm, open label randomized controlled interventional clinical trial to assess the effect of the BAF inhibitor pyrimethamine on the HIV-1 reservoir when given alone or as a combination of LRAs with the histone deacetylase inhibitor valproic acid. Individuals aged ≥18 years visiting the outpatient department of infectious diseases at the Erasmus University Medical Center, Rotterdam, the Netherlands were eligible if they had a confirmed HIV-1 infection and a CD4+ T cell count of at least 200 cells/μl, receiving cART with a pre-cART HIV-1 RNA zenith of ≥10 000 copies/mL and plasma HIV-1 RNA levels of <20 copies/mL in the year prior to the intervention (with a minimum of two measurements taken). Exclusion criteria were female with a reproductive potential due to valproic acid’s teratogenic potential, previous virological failure with resistance-associated mutations acquired on cART, active hepatitis B or C infection, prior exposure to LRAs, immunomodulating medication or medication known to interact with valproic acid or pyrimethamine. The inclusion period of the study was between April 2018 and September 2020 following approval by the Medical Research Ethics Committee (MEC-2017-476) in accordance with the principles of the Helsinki Declaration. The protocol was amended once in February 2020 before the analysis of endpoints. The reason for this amendment was to allow additional blood sampling to overcome potential low cell yields for a main endpoint analysis on the reservoir size. All participants provided written informed consent and participant/sample identifiers included were not known to anyone (e.g. participants themselves or hospital staff) outside the research group. The full protocol is available as appendix and a synopsis is available in the supplementary data. Study reporting follows the CONSORT reporting guidelines.

### Intervention

Eligible and consenting trial participants were screened within six weeks before the start of the intervention phase, and subsequently randomized to one of four arms to receive oral doses of either: pyrimethamine once a day for 14 days (200 mg on the first day followed by 100 mg onwards); 30 mg/kg/day valproic acid (divided into two equal doses) for 14 days; a combination of pyrimethamine and valproic acid dosed likewise; or no intervention. For the randomization process, an independent statistician delivered sealed opaque envelopes that included the study identifier with the allocated treatment based on random allocation performed in R. Valproic acid and pyrimethamine were administered in dosages used in the chronic treatment of epilepsy,(*60*) and previous clinical studies on the treatment of cerebral toxoplasmosis in AIDS patients or as prophylaxis or malarial treatment in pregnant women (*49, 53–57*). The dose and duration of valproic acid and pyrimethamine treatment, and the timing of viral reservoir analysis on day 0 after 6 hours was based on the T_max_ of both compounds, available literature, expert opinion and our previous work (*67–69*). Blood samples were collected 6 hours after the participants received the intervention regimen assuming the T_max_ to be reached by pyrimethamine and valproic acid after approximately 4 hours, and an additional 2-3 hours for HIV-1 transcription and HIV-1 RNA accumulation (*24*). To achieve a constant and the earliest T_max_ possible, participants received their first dose of the intervention regimen on day 0 in a fasted state. On day 0, cART and study medication intake were taken under direct supervision of the study staff. The two-week intervention phase was followed by a four-week post intervention phase. Overall, participation involved 11 study visits. In order to obtain an additional post intervention phase measurement for the inducible HIV-1 reservoir size, the study was amended and participants were asked to give written consent to an extra blood draw after the four-week post-intervention phase ended. Plasma HIV-1 RNA levels (COBAS TaqMAN; Roche, Basel, Switzerland; limit of quantification 20 copies/mL) were monitored at all 11 visits. CD4+ T cell counts were determined at screening, at the start and end of the intervention phase (day 0 and day 14), at day 42, and at the post intervention phase (≥ day 42).

### Safety

Hematological parameters, kidney and liver function were monitored pretreatment, day 7, day 14 and day 42. At each study visit, patients were clinically assessed by the study physician and safety assessments were performed. All patient-reported adverse events (AE) and serious adverse events (SAE) were evaluated in relation to valproic acid and pyrimethamine. Severity was graded according to the Common Terminology Criteria for Adverse Events (CTCAE) version 4.0. In the study protocol, a number of stopping rules were defined including a pre-specified interim analysis after the inclusion of 14 participants focusing on the number of participants discontinuing study medication during the intervention phase, CD4+ T cell count <200 cells/μl, possibly drug related SAE/AE ≥ grade 4, or AIDS related illness (CDC-C events).

### Study endpoints

The pre-specified primary endpoint was the change in HIV-1 reactivation at treatment initiation and at the end of treatment in the study arms, measured as the change in cell associated unspliced HIV-1 RNA (CA US HIV-1 RNA) between treatment initiation (day 0 at 0 hours) and at the end of the study (day 42). Secondary endpoints reported here included the change in inducible HIV-1 reservoir size as quantified by tat/rev induced limiting dilution assay (TILDA), the synergistic effects of pyrimethamine with valproic acid on the induction of CA US HIV-1 RNA, the change in CA US HIV-1 RNA between timepoints within and between study arms, plasma HIV-1 RNA changes, the pharmacokinetic and pharmacodynamic profiles of the intervention regimens, and the clinical safety and tolerability of the intervention regimen.

### Cell associated unspliced HIV-1 RNA

For quantification of CA US HIV-1 RNA, CD4+ T cells were isolated from peripheral blood mononuclear cells (PBMCs) by negative selection using EasySep Human CD4+ T Cell Enrichment Cocktail (StemCell Technologies). Isolation of CD4+ T cells was performed on ice or at 4 °C (unless indicated otherwise by the manufacturer). Approximately 1.5 x 10^6^ CD4+ T cells were lysed in triplicate with TRI reagent. Total RNA was isolated by phenol/chloroform isolation method following manufacturer’s instructions. A minimum of 150 ng of Total RNA was DNAse-treated following manufacturer’s instructions (DNAseI Amplification Grade Invitrogen) and complementary DNA (cDNA) synthesis was performed in duplicate with SuperScript II (Invitrogen) following manufacturer ‘s instructions. Absolute quantification of CA US HIV-1 RNA was performed following a modified version of Pasternak *et al.* methodology (*70*). Briefly, first round of nested amplification was performed in a final volume of 25 μL using 10 μL of cDNA, 2.5 μL of 10x PCR buffer (Life Technologies), 1 μL of 50 mM MgCl2 (Life Technologies), 1 μL of 10 mm dNTPs (Life Technologies, 0.075 μL of 100 μM US Forward primer, 0.075 μL of 100 μM US Reverse 1 primer, and 0.2 μL Platinum Taq polymerase (Life Technologies) at 95°C for 5 min, followed by 15 cycles at 95°C for 30 seconds, 55°C for 30 seconds and 72°C for 15 min. The second round of amplification was performed in a final volume of 25 µL using 2 µL of pre-amplified cDNA, 2.5 μL of 10x PCR buffer (Life Technologies), 1 μL of 50 mM MgCl2 (Life Technologies), 1 μL of 10 mm dNTPs (Life Technologies), 0.05 μL of 100 μM US Forward primer, 0.05 μL of 100 μM US Reverse 2 primer, 0.0375 μL of US Probe and 0.2 μL Platinum Taq polymerase (Life Technologies) at 95°C for 5 min, followed by 45 cycles at 95°C for 30 seconds and 60°C for 1 min. List of primers and probe is available in **Table S8**. Absolute number of US copies in the PCR was calculated using a standard curve ranging from 2 to 512 copies of a plasmid containing the full-length HIV-1 genome (pNL4.3.Luc.R-E-). Based on standard curve analysis we assigned a limit of quantification (LOQ) of 16 copies with an intra assay coefficient of variation of <5% (**Figure S4D**). The quantity of CA US HIV-1 RNA was expressed as number of copies per 150 ng of input RNA in reverse transcription.

### BAF complex target gene expression

RT-qPCR reactions were conducted using GoTaq qPCR Master Mix (Promega) following the manufacturer’s protocol. The following thermal cycling protocol was used for amplification: 3 min at 95 °C, followed by 40 cycles of 95 °C for 10 s and 60 °C for 30 s. Expression data was calculated using 2-ddCt methodology (*71*). Cyclophilin A was used as housekeeping gene for the analysis. List of primers is available in **Table S8**.

### Pharmacokinetical analysis

Self-reported adherence was assessed at each study visit and empty pill strips were collected after the two-week intervention phase. EDTA plasma samples were collected at the start of the intervention phase on day 0 (at 0 hours and after 6 hours post first dose), day 7 (valproic acid only), day 14, day 42. Plasma concentrations of valproic acid were analyzed by using a routinely implemented and validated assay (Multigent® Valproic Acid Assays) at the Erasmus University Medical Center Pharmacy. For pyrimethamine measurements, we developed an assay (validated according to FDA/EMA guidelines) using the Waters Acquity UPLC-MS/MS. Samples (50µL) were prepared and pumped into the column (Waters Acquity BEH C18 (1.7 µm, 2.1 x 100 mm), at 50 ℃). A 0.3 ml/min gradient with two eluents was used (A: 2 mM ammonium acetate + 0.1% formic acid in water; B: 2 mM ammonium acetate + 0.1% formic acid in methanol). The total run time was 4.2 min. Mass transitions used were m/z 249.09 to 176.97 (pyrimethamine) and m/z 325.05 to 307.05 (quinine), cone voltages were 22 V (pyrimethamine) and 58 V (quinine), collision energy were 20 eV (pyrimethamine) and 14 eV (quinine). Capillary voltage was 3 kV, source temperature 150 ℃, desolvation temperature 400℃, desolvation gas flow 900 L/h. For measurement validation, we performed linearity, correctness, limit of detection (LOD) and lower limit of quantification (LLOQ), repeatability, reproducibility, measurement uncertainty, robustness, and carryover. The detection range was validated between 0.3 and 22 mg/L. Unbound dolutegravir plasma concentrations were quantified with a validated UPLC-MS/MS bioquantification method as previously described (*59*).

### Inducible HIV-1 reservoir

The frequency of CD4+ T cells expressing multiply spliced (MS) HIV-1 RNA was determined using the tat/rev induced limiting dilution assay (TILDA) with some modifications to the protocol described previously by our group (*66*). Briefly, total CD4+ T cells were isolated from PBMCs by negative magnetic selection using EasySep CD4+ Human CD4+ T Cell Enrichment kit (StemCell Technologies). Following isolation, 1.5 x 10^6^ CD4+ T cells/mL were rested in complete RPMI-1640 for 5-8 hours prior to 12 hours of stimulation with 100 ng/mL phorbol 12-myristate 13-acetate (PMA) and 1 μg/mL ionomycin (both from Sigma Aldrich). After stimulation, CD4+ T cells were washed and resuspended in serum-free RPMI-1640. Cells were counted and serially diluted accordingly; 1.8 x 10^6^ cells/mL, 9 x 10^5^ cells/mL, 3 x 10^5^ cells/mL, 1x 10^5^ cells/mL. For certain samples with smaller reservoirs, TILDA was performed using a higher input of CD4+ T cells (2 to 4-fold). In the pre-amplification reaction, 10 μL of the cell suspension from each dilution was dispensed into 24-48 wells of a 96 well plate containing 2 μL One-step RT-PCR enzyme (Qiagen), 10 μL 5x One-step RT-PCR buffer (Qiagen), 10 μL Triton X-100 (0.3 %), 0.25 μL RNAsin (40U/μL), 2 μL dNTPs (10 mM each), 1 μL tat1.4 forward primer (20 μM) and rev reverse primer (20 μM) (as published), and nuclease-free water to a final reaction volume of 50 μL. The one-step RT-PCR was run using the following thermocycling conditions: 50 °C for 30 min, 95 °C for 15 min, followed by 25 cycles of 95 °C for 1 min, 55 °C for 1 min and 72 °C for 2 min, and a final extension at 72 °C for 5 min. After pre-amplification, 2 μL of the products were used as input for the real time PCR to detect tat/rev MS HIV-1 RNA in a final reaction volume of 20 μL, which consisted of 5 μL 4x Taqman Fast Advanced Master mix (Thermo Fisher Scientific), 0.4 μL of tat2.0 forward primer, rev reverse primer (each at 20 μM), and MS *tat/rev* probe (5 μM). The real time PCR was performed using the following programme: 50 °C for 5 min, 95 °C for 20 s, followed by 45 cycles of 95 °C for 3 s, 60 °C 30 s. Positive wells at each dilution were recorded and used to determine the frequency of cells expressing tat/rev MS HIV-1 RNA by maximum likelihood method. The inter-operator reproducibility of the assay was evaluated using samples obtained from participants in the pyrimethamine arm (**Supp. Fig.5B-D**). Primers used for qPCR to generate the endpoints by TILDA and the other PCR based assays are listed in supplementary table S8.

### Statistical analysis

The sample size was based on the null hypothesis that the change in CA US HIV-1 RNA between the study arms are equal and the alternative hypothesis that the change in CA US HIV-1 RNA are not equal. Assuming a mean change of 31 copies CA US HIV-1 RNA (standard deviation estimated at 20) by the intervention based on previous trials,(*35*) we could detect a twofold increase between any of the 4 groups with 6 patients per group at 80% power and alpha 0.05. A twofold change has also been observed in other clinical trials and in our preclinical experiments with pyrimethamine ex vivo (*24, 34, 44*). This therefore was considered a realistic target to substantiate the power calculation on. We included 7 patients per study arm to account for drop out.

The measurements related to the endpoints CA US HIV-1 RNA, multiply spliced HIV-1 RNA, plasma HIV-1 RNA and drug plasma levels were described as median with interquartile (IQR) or full range. When the CA US HIV-1 RNA measurement was below the limit of quantitation set at 16 copies/150ng total RNA, we imputed 16 copies at these datapoints for further calculations. The fold change difference per study arm from pretreatment CA US HIV-1 RNA levels to the timepoints 6 hours, day 14 and day 42 after first dosing were calculated by adding up the fold change per individual divided by the total number of measurements available per timepoint. We analysed the primary endpoint by a generalized estimating equation (GEE) model to evaluate whether any difference in the CA US HIV-1 RNA existed at any timepoint during the trial in any of the 3 interventional arms compared to the control arm. An interaction term of time and treatment was included. We used the Bliss independence method to conclude on synergism between pyrimethamine and valproic acid using the following equation μ_(1+2)exp._ = [1 − (1 – μ_1_) × (1− μ_2_)]. The difference between the observed and expected amount of CA US HIV-1 RNA was calculated and combination therapy was considered synergistic if the difference and its 95% confidence interval were >0. Due to the number of patients in this pilot, all secondary endpoints were exploratory. We therefore limited the use of inferential statistics to assessing the median fold changes between the timepoints in CA US HIV-1 RNA within and between treatment arms by Wilcoxon signed rank tests, Mann Whitney U tests and we used Pearson test to explore correlations between clinical variables and plasma pyrimethamine levels with the primary endpoint CA US HIV-1 RNA. We did not adjust for multiple testing.

## Data Availability

All data, code, and materials used in the analysis are available to any researcher for purposes of reproducing or extending the analysis upon request and approval by both corresponding authors.

## ACKNOWLEDGEMENTS

We want to express our gratitude to all participants in the study. Furthermore, we want to thank all internist-infectious diseases specialists at Erasmus MC for their help in recruitment and the scientists from the Erasmus MC HIV Eradication Group (EHEG). Pauline Bollen is acknowledged for her help with analyzing the dolutegravir-valproic acid interaction. The logistics and sample processing of the work were greatly supported by the research nurses (Rene van Engen, Ayten Karisli, Maartje Wagemaker), laboratory technicians, and the Msc students who contributed (Katie Hensley, Daniek Teijema). Lastly, this study would not have been possible without the scientific vision of Charles A.B. Boucher, a professor in virology and our dear colleague, who unfortunately passed away too early in February 2021.

## Author contributions

Conceptualization: HABP, RG, AV, TM, CR

Methodology: HABP, RC, CL, SR, LL, RO, TH, JVK, AC, DB, RJP, BCP, RG, YMM, PDK, AV, TM, CR

Investigation: HABP, RC, CL, SR, LL, TH, EvN, MdMM, HB, TdVS, JVK, AC, DB, ThM, RJP, BCP, RG, YMM, PDK, AV, TM, CR

Visualization: HABP, RC, CL, TM, CR

Funding acquisition: SR, RG, AV, TM, CR

Project administration: HABP, RC, CL, TM, CR

Supervision: SR, DB, RJP, BCP, RG, PDK, AV, TM, CR

Writing – original draft: HABP, RC, CL, TM, CR

Writing – review & editing: HABP, RC, CL, SR, LL, RO, TH, EvN, MdMM, HB, TdVS, JVK, AC, DB, ThM, RJP, BCP, RG, YMM, PDK, AV, TM, CR

## SUPPLEMENTARY DATA

**Fig. S1.**
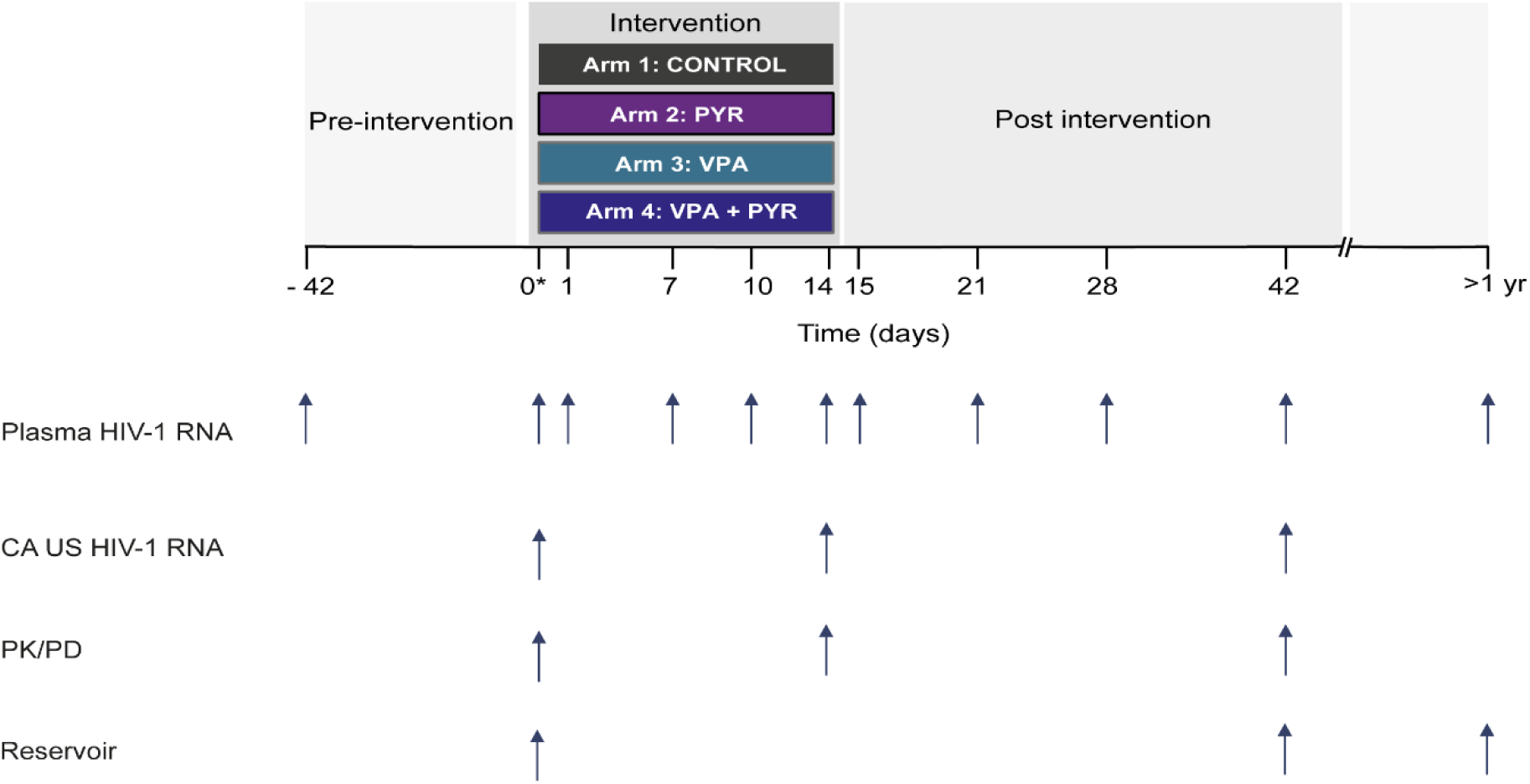
Study design overview. The screening time point is shown at day −42. The three treatment arms and a fourth control arm are shown in blue (VPA arm), purple (PYR arm), dark blue (VPA+PYR), and black (control). Sampling time points for primary and secondary endpoint analysis are indicated by small solid arrows below the timeline. All participants provided 1 extra blood sample at least 1 year after the day 42 visit. * On day 0, CA HIV-1 RNA was measured in samples taken before drug intake (t=0 hours) and after 6 hours. VPA: valproic acid; PYR: pyrimethamine; CA HIV-1 RNA: cell-associated HIV-1 RNA; PK: pharmacokinetics; PD: pharmacodynamics.

**Fig. S2.**
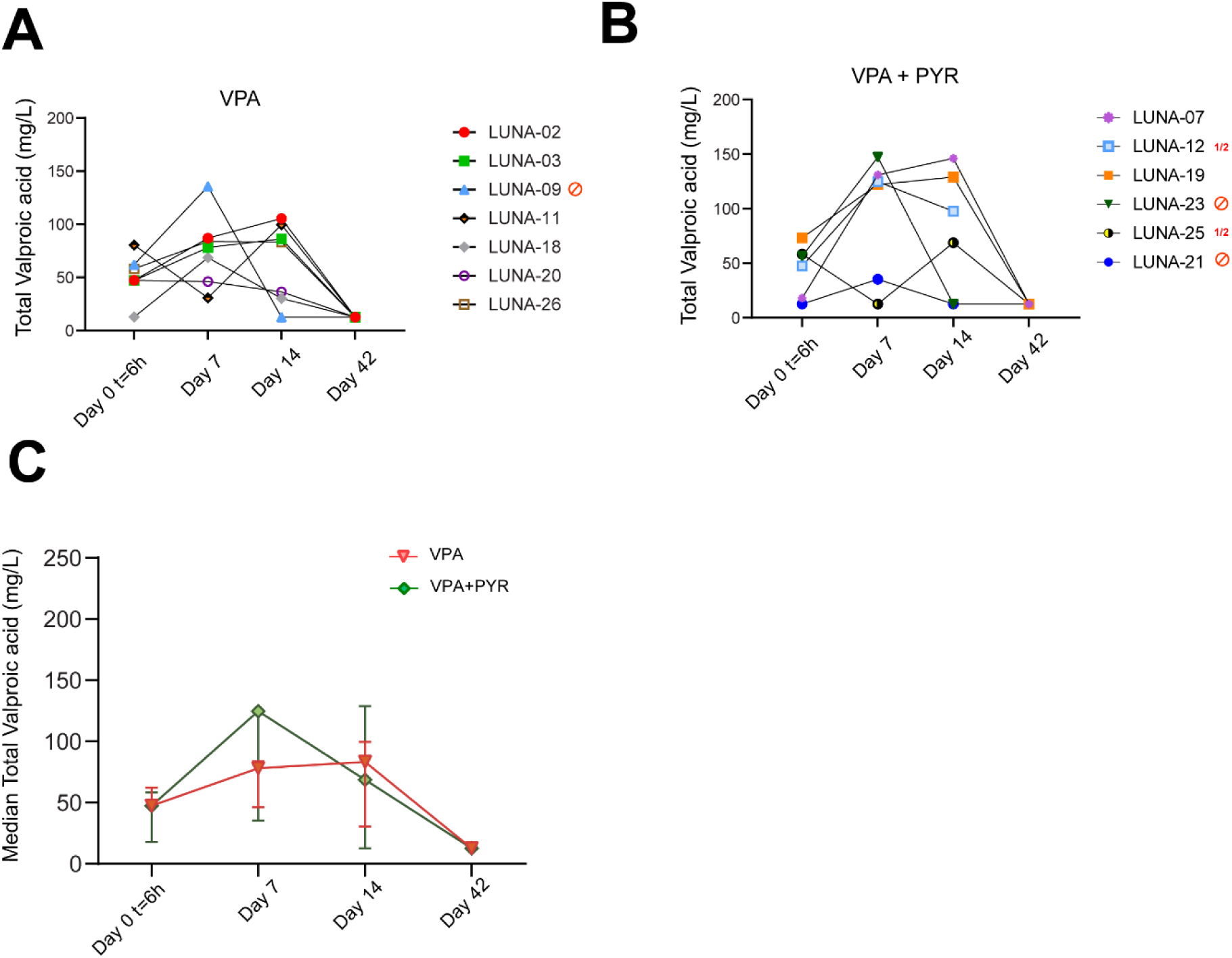
Pharmacokinetics of the drug valproic acid in study participants. (A-B) Pharmacokinetics analysis of the drug valproic acid in study participants represented as plasma valproic acid levels per individual on valproic acid (A) or the combination treatment with pyrimethamine and valproic acid (B) and the median total valproic acid (mg/L) level in plasma (C) including all participants of the valproic acid arm (red) and the combination treatment arm with pyrimethamine and valproic acid (green) at 6 hours after first dosing (day 0, t=6hr), at day 7, at the end of treatment period (day 14) and 28 days after end of treatment (day 42). Valproic acid concentration measurement was unsuccessful at day 42 for LUNA-12. Participants that stopped study medication or had dose adjustments are indicated with either a red stop sign or red ½ symbol and a red cross on the individual points in the graph. In the valproic acid arm one participant stopped his study medication on day 8 (LUNA-09). In the combination treatment arm, three participants stopped both compounds (LUNA-23 on day 3, LUNA-06 on day 7 and LUNA-21 on day 10), and two participants had their dosage adjusted (LUNA-25 had VPA dose halved from day 2 on, PYR dose halved from day 7 on, and LUNA-12 had PYR dose halved from day 7 on).

**Fig. S3.**
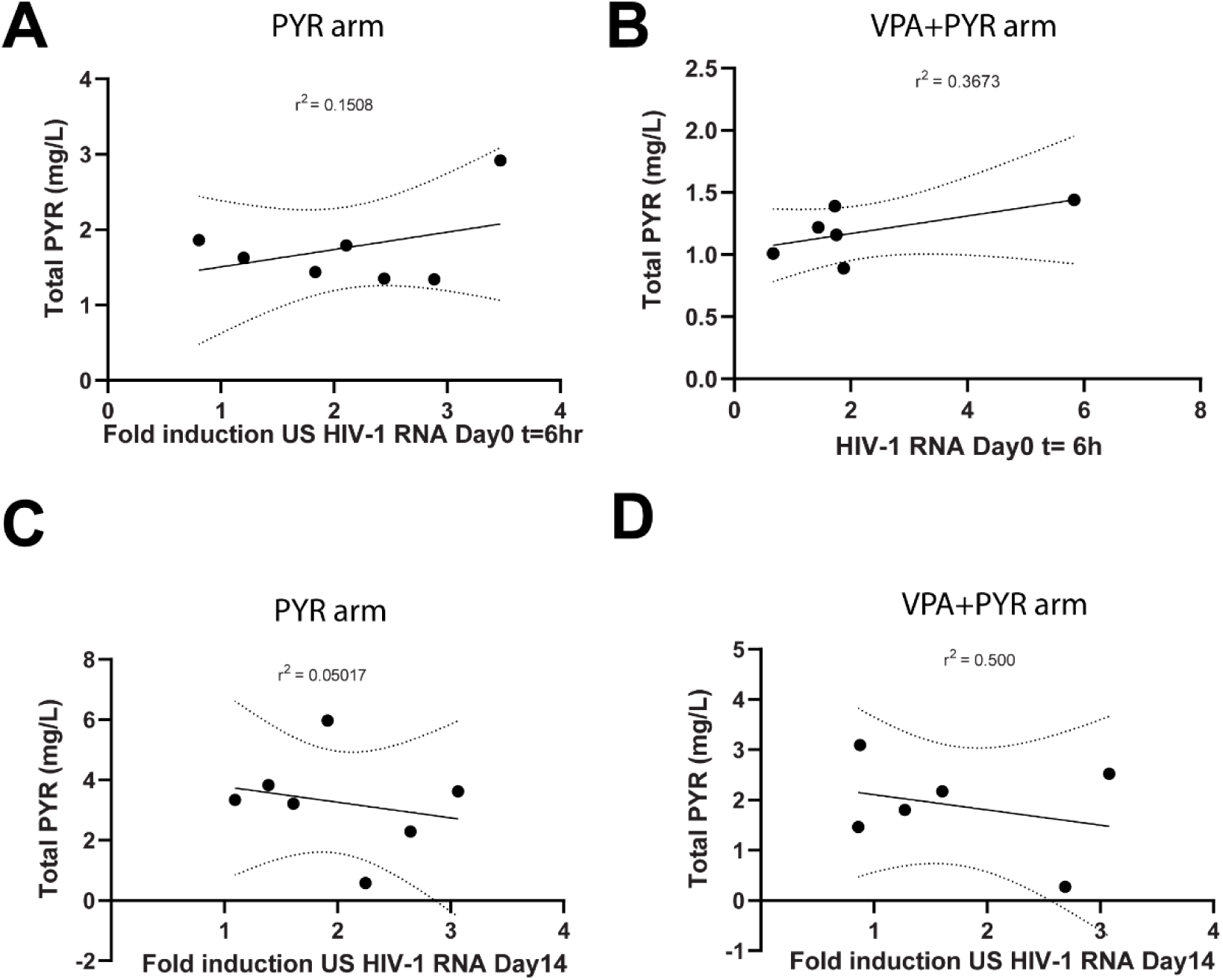
Correlation plots of pyrimethamine plasma levels with the primary endpoint cell associated HIV-1 RNA. Mean and 95% confidence interval correlations with coefficient values (r) between total pyrimethamine (mg/L) and CA US HIV-1 RNA fold change at 6 hours after the first dosing participants on pyrimethamine (A), combination treatment with pyrimethamine and valproic acid (B) and at d14 for these 2 groups (C-D) in 13 of 14 participants exposed to pyrimethamine with sufficient cells available for CA US HIV-1 RNA measurements. Coefficients were calculated by Pearson correlations.

**Fig. S4.**
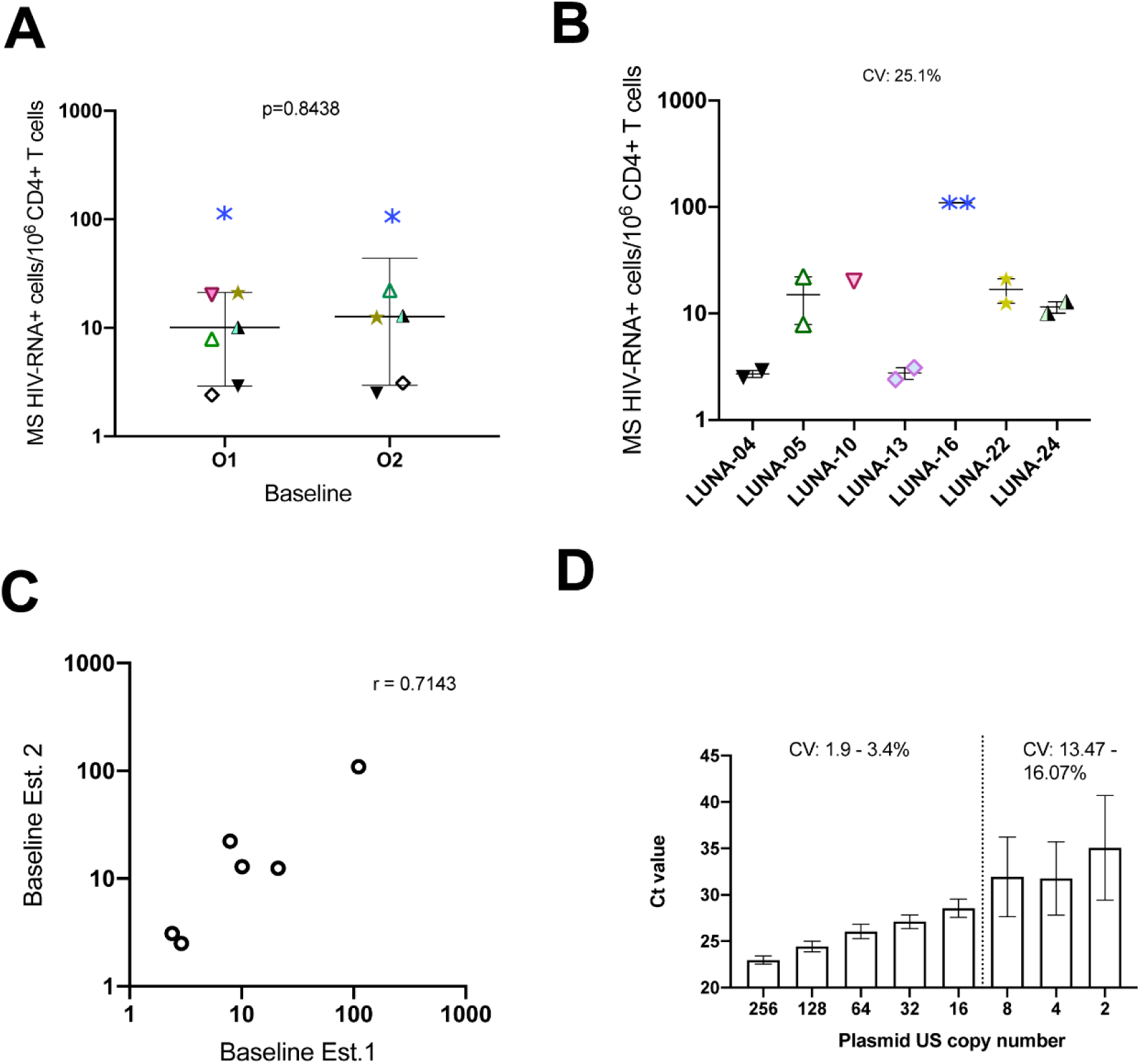
Assay reproducibility and standard curve analysis. (A) and (B) Inter-operator reproducibility of TILDA. Measurements of inducible reservoir size in baseline samples obtained from participants in the pyrimethamine arm. TILDA was performed by different operators [O1 and O2] in independent experiments. The error bars represent the interquartile range. A Wilcoxon matched-pairs signed rank test was used to compare the medians of the TILDA data generated by O1 and O2. CV indicates coefficient of variation. (C) Spearman correlation of the TILDA data between operators. (D) Standard curve interpolation of absolute number of US copies ranging from 2 to 256 copies of a plasmid containing the full-length HIV-1 genome (pNL4.3.Luc.R-E-) and cycle threshold value (Ct value) in the PCR. CV indicates coefficient of variation. Based on standard curve analysis we assigned a limit of quantification (LOQ) of 16 copies with an intra assay coefficient of variation of <5%.

**Table S1.** Total number of self-reported adverse events and their severity during trial participation.

|  | **Grade 1** | **Grade 2** | **Grade3** | **Any Grade** |
| --- | --- | --- | --- | --- |
| Control arm | 4 | 0 | 0 | 4 |
| Pyrimethamine arm | 33 | 3 | 2 | 38 |
| Valproic acid arm | 13 | 2 | 0 | 15 |
| Valproic acid + pyrimethamine arm | 32 | 13 | 1 | 46 |
| **Total adverse events** | **82** | **18** | **3** | **103** |
| Presumed related | 72 | 16 | 3 | 91 |
| Presumed not related | 10 | 2 | 0 | 12 |

**Table S2.**
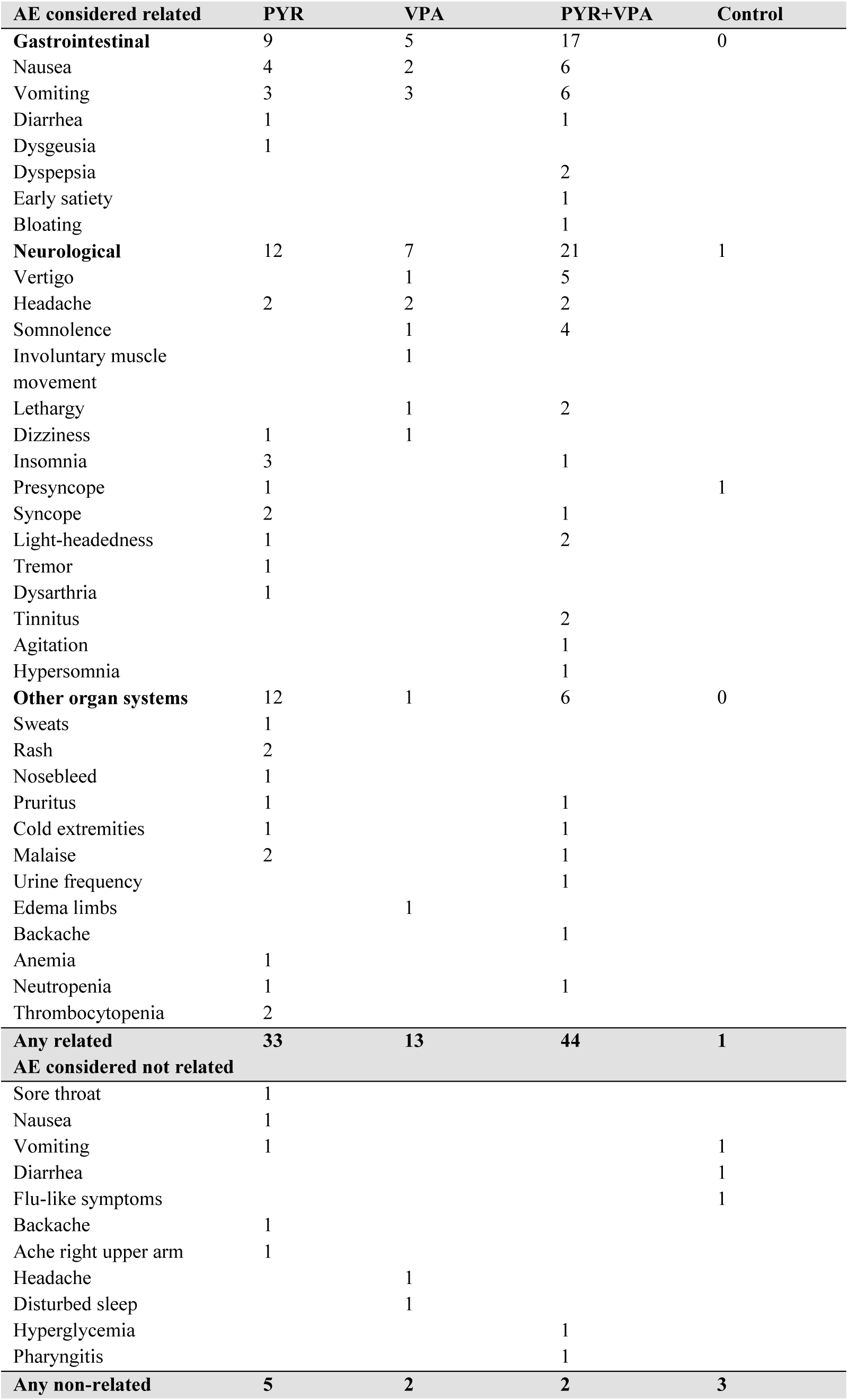
Total number of self-reported adverse events (AE) per treatment arm during trial participation.

| **AE considered related** | **PYR** | **VPA** | **PYR+VPA** | **Control** |
| --- | --- | --- | --- | --- |
| **Gastrointestinal** | 9 | 5 | 17 | 0 |
| Nausea | 4 | 2 | 6 |  |
| Vomiting | 3 | 3 | 6 |  |
| Diarrhea | 1 |  | 1 |  |
| Dysgeusia | 1 |  |  |  |
| Dyspepsia |  |  | 2 |  |
| Early satiety |  |  | 1 |  |
| Bloating |  |  | 1 |  |
| **Neurological** | 12 | 7 | 21 | 1 |
| Vertigo |  | 1 | 5 |  |
| Headache | 2 | 2 | 2 |  |
| Somnolence |  | 1 | 4 |  |
| Involuntary muscle movement |  | 1 |  |  |
| Lethargy |  | 1 | 2 |  |
| Dizziness | 1 | 1 |  |  |
| Insomnia | 3 |  | 1 |  |
| Presyncope | 1 |  |  | 1 |
| Syncope | 2 |  | 1 |  |
| Light-headedness | 1 |  | 2 |  |
| Tremor | 1 |  |  |  |
| Dysarthria | 1 |  |  |  |
| Tinnitus |  |  | 2 |  |
| Agitation |  |  | 1 |  |
| Hypersomnia |  |  | 1 |  |
| **Other organ systems** | 12 | 1 | 6 | 0 |
| Sweats | 1 |  |  |  |
| Rash | 2 |  |  |  |
| Nosebleed | 1 |  |  |  |
| Pruritus | 1 |  | 1 |  |
| Cold extremities | 1 |  | 1 |  |
| Malaise | 2 |  | 1 |  |
| Urine frequency |  |  | 1 |  |
| Edema limbs |  | 1 |  |  |
| Backache |  |  | 1 |  |
| Anemia | 1 |  |  |  |
| Neutropenia | 1 |  | 1 |  |
| Thrombocytopenia | 2 |  |  |  |
| **Any related** | **33** | **13** | **44** | **1** |
| **AE considered not related** |  |  |  |  |
| Sore throat | 1 |  |  |  |
| Nausea | 1 |  |  |  |
| Vomiting | 1 |  |  | 1 |
| Diarrhea |  |  |  | 1 |
| Flu-like symptoms |  |  |  | 1 |
| Backache | 1 |  |  |  |
| Ache right upper arm | 1 |  |  |  |
| Headache |  | 1 |  |  |
| Disturbed sleep |  | 1 |  |  |
| Hyperglycemia |  |  | 1 |  |
| Pharyngitis |  |  | 1 |  |
| **Any non-related** | **5** | **2** | **2** | **3** |

**Table S3.** Total number of self-reported adverse events (AE) and their severity during trial participation.

| **AE considered related** | **Grade 1**<br>**N(%)** | **Grade 2**<br>**N(%)** | **Grade3**<br>**N(%)** | **Any Grade**<br>**N(%)** | **No. of patients**<br>**N(%)** |
| --- | --- | --- | --- | --- | --- |
| Malaise | 2 | 1 | 0 | 3 | 3 |
| Nausea | 8 | 4 | 0 | 12 | 12 |
| Dyspepsia | 2 | 0 | 0 | 2 | 2 |
| Early satiety | 1 | 0 | 0 | 1 | 1 |
| Bloating | 1 | 0 | 0 | 1 | 1 |
| Vomiting | 11 | 1 | 0 | 12 | 9 |
| Diarrhea | 2 | 0 | 0 | 2 | 2 |
| Headache | 5 | 1 | 0 | 6 | 6 |
| Somnolence | 5 | 0 | 0 | 5 | 4 |
| Hypersomnia | 1 | 0 | 0 | 1 | 1 |
| Insomnia | 4 | 0 | 0 | 4 | 4 |
| Involuntary muscle movement | 1 | 0 | 0 | 1 | 1 |
| Vertigo | 3 | 3 | 0 | 6 | 5 |
| Tinnitus | 1 | 1 | 0 | 2 | 2 |
| Lethargy | 2 | 1 | 0 | 3 | 3 |
| Agitation | 0 | 1 | 0 | 1 | 1 |
| Dizziness | 2 | 0 | 0 | 2 | 2 |
| Light headedness | 3 | 0 | 0 | 3 | 3 |
| Presyncope | 2 | 0 | 0 | 2 | 2 |
| Syncope | 0 | 0 | 3 | 3 | 3 |
| Edema limbs | 1 | 0 | 0 | 1 | 1 |
| Cold extremities | 2 | 0 | 0 | 2 | 2 |
| Sweats | 1 | 0 | 0 | 1 | 1 |
| Rash | 2 | 0 | 0 | 2 | 1 |
| Pruritus | 2 | 0 | 0 | 2 | 2 |
| Dysarthria | 1 | 0 | 0 | 1 | 1 |
| Dysgeusia | 1 | 0 | 0 | 1 | 1 |
| Tremor | 1 | 0 | 0 | 1 | 1 |
| Urinary frequency | 1 | 0 | 0 | 1 | 1 |
| Back pain | 1 | 0 | 0 | 1 | 1 |
| Nosebleed | 1 | 0 | 0 | 1 | 1 |
| Anemia | 1 | 0 | 0 | 1 | 1 |
| Platelet count decreased | 1 | 1 | 0 | 2 | 1 |
| Neutrophil count decreased | 0 | 2 | 0 | 2 | 2 |
| **Any related AE** | **72** | **16** | **3** | **91** | **84** |
| **AE considered not related** |  |  |  |  |  |
| Nausea | 1 | 0 | 0 | 1 | 1 |
| Vomiting | 2 | 0 | 0 | 2 | 2 |
| Diarrhea | 1 | 0 | 0 | 1 | 1 |
| Headache | 1 | 0 | 0 | 1 | 1 |
| Disturbed sleep | 1 | 0 | 0 | 1 | 1 |
| Ache right upper arm | 0 | 1 | 0 | 1 | 1 |
| Upper back ache | 1 | 0 | 0 | 1 | 1 |
| Sore throat | 1 | 0 | 0 | 1 | 1 |
| Hyperglycemia | 0 | 1 | 0 | 1 | 1 |
| Pharyngitis | 1 | 0 | 0 | 1 | 1 |
| Flu like symptoms | 1 | 0 | 0 | 1 | 1 |
| Any not related | 10 | 2 | 0 | 12 | 12 |
| **Total AE** | **82** | **18** | **3** | **103** | **96** |

**Table S4.**
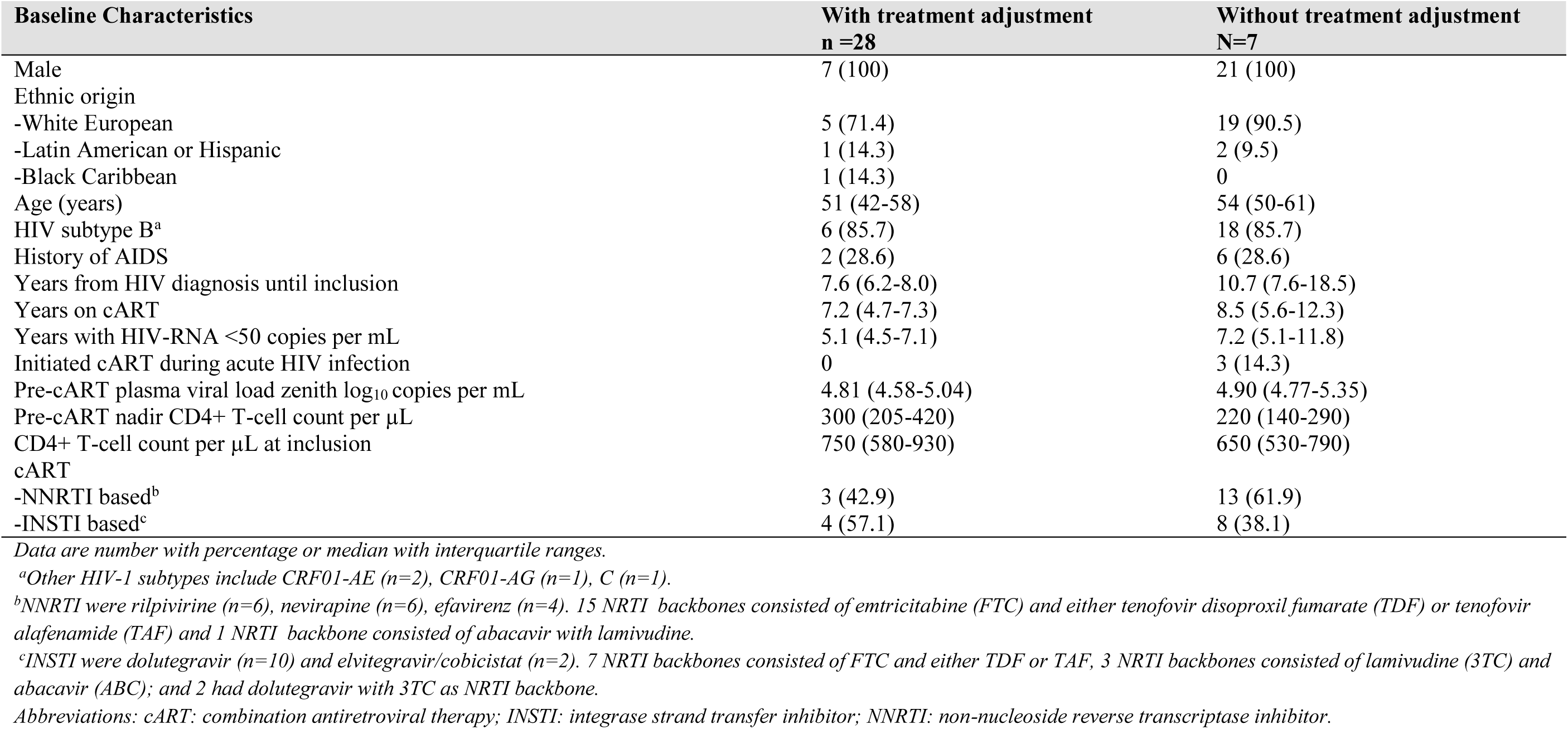
Baseline characteristics of participants with and without adjusted interventions.

**Table S5.** Individual clinical characteristics and cell associated unspliced HIV-1 RNA during LUNA.

|  | CD4 T-cell<br>count per<br>µL at<br>inclusion | Years on<br>cART | cART | CA US<br>HIVRNA<br>copies/150ng<br>Total RNA<br>Day 0,<br>pretreatment | CA US<br>HIVRNA<br>copies/150ng<br>Total RNA Day<br>0,<br>6 hours | CA US<br>HIVRNA<br>copies/150ng<br>Total RNA<br>Day 14 | CA US<br>HIVRNA<br>copies/150ng<br>Total RNA<br>Day 42 |
| --- | --- | --- | --- | --- | --- | --- | --- |
| **PYR** |  |  |  |  |  |  |  |
| LUNA-04 | 580 | 7,3 | FTC, TAF, EVG/c | 32,60 | 94,04 | 62,36 | 101,16 |
| LUNA-05 | 390 | 5,5 | FTC, TDF, EFV | 79,98 | 96,01 | 111,26 | 84,39 |
| LUNA-10 | 810 | 7,2 | FTC, TAF, NVP | 51,15 | 125,01 | 114,91 | 85,80 |
| LUNA-13 | 850 | 5,6 | FTC, TDF, EFV | 68,92 | 126,50 | 111,02 | 54,17 |
| LUNA-16 | 400 | 16,3 | FTC, TDF, NVP | 24,35 | 51,31 | 74,67 | 82,71 |
| LUNA-22 | 610 | 7,7 | FTC, TAF, DTG | 49,43 | 39,82 | 54,10 | 69,81 |
| LUNA-24 | 730 | 23,1 | FTC, TAF, NVP | 16,00 | 55,52 | 42,33 | 29,28 |
| **PYR+VPA** |  |  |  |  |  |  |  |
| LUNA-06<sup>1</sup> | 630 | 7,2 | FTC, TAF, RPV |  |  |  |  |
| LUNA-07 | 960 | 12,2 | FTC, TDF, RPV | 53,68 | 35,85 | 46,27 | 28,10 |
| LUNA-12 | 1050 | 3,5 | 3TC, ABC, DTG | 34,27 | 49,48 | 54,91 |  |
| LUNA-19 | 240 | 5,6 | FTC, TAF, RPV | 32,97 | 57,84 | 28,92 |  |
| LUNA-21 | 420 | 4,8 | FTC, TDF, RPV | 28,25 | 48,73 | 35,93 | 45,78 |
| LUNA-23 | 530 | 7,5 | 3TC, ABC, DTG | 16,64 | 97,05 | 51,25 | 49,58 |
| LUNA-25 | 1180 | 4,5 | 3TC, DTG | 21,83 | 41,01 | 58,74 | 31,47 |
| **VPA** |  |  |  |  |  |  |  |
| LUNA-02 | 790 | 4,8 | FTC, TAF, RPV | 95,16 | 101,51 | 161,79 | 59,33 |
| LUNA-03 | 590 | 7,6 | FTC, TAF, DTG | 12,93 | 10,16 | 18,41 | 16,00 |
| LUNA-09 | 750 | 7,4 | FTC, TAF, DTG | 22,90 | 42,74 | 28,40 | 49,74 |
| LUNA-11 | 1250 | 21,1 | FTC, TDF, NVP | 39,10 | 53,04 | 97,80 | 59,03 |
| LUNA-18 | 530 | 12,3 | FTC, TDF, NVP | 16,00 | 19,23 | 16,00 | 16,00 |
| LUNA-20 | 830 | 4,3 | FTC, TDF, EFV | 76,70 | 51,59 | 47,89 | 52,98 |
| LUNA-26 | 330 | 9,3 | FTC, TAF, DTG | 16,00 | 16,00 | 18,82 | 22,83 |
| **CONTROL** |  |  |  |  |  |  |  |
| LUNA-01 | 780 | 3,9 | FTC, TDF, RPV | 16,00 | 16,00 | 16,00 | 16,00 |
| LUNA-08 | 700 | 14,7 | FTC, TDF, EFV | 50,82 | 63,21 | 57,75 | 45,16 |
| LUNA-14 | 650 | 7,4 | 3TC, DTG | 120,00 | 114,52 | 93,02 | 100,41 |
| LUNA-15 | 680 | 21,7 | FTC, TAF, EVG/c | 62,96 | 113,72 | 113,54 | 120,88 |
| LUNA-17 | 530 | 8,5 | 3TC, ABC, NVP | 16,00 | 39,37 | 29,24 | 22,56 |
| LUNA-27 | 980 | 9,8 | FTC, TDF, DTG | 16,00 | 16,00 | 16,00 | 16,00 |
| LUNA-28 | 470 | 11,2 | 3TC, ABC, DTG | 16,00 | 16,00 | 16,00 | 16,00 |

<sup>1</sup> LUNA-06 had insufficient PBMC at day 0 at 6 hours post dosing to measure the primary endpoint. Due to the missing baseline measurement, no CA US HIV-1 RNA were measured at the other timepoints. *Abbreviations: cART: combination antiretroviral therapy; PYR: pyrimethamine; VPA: valproic acid; FTC: emtricitabine; 3TC: lamivudine; ABC: abacavir; TDF: tenofovir disoproxil fumarate; TAF: tenofovir alafenamide; EFV: efavirenz; NVP: nevirapine; RPV: rilpivirine; DTG: dolutegravir; EVG/c: elvitegravir/cobicistat; .*

**Table S6:** plasma HIV-1 RNA evolution during LUNA per participant.

|  | Day<br>0, 0hr | Day<br>0, 6hr | Day<br>1 | Day<br>7 | Day<br>10 | Day<br>14 | Day<br>15 | Day<br>21 | Day<br>28 | Day<br>42 |
| --- | --- | --- | --- | --- | --- | --- | --- | --- | --- | --- |
| **PYR** |  |  |  |  |  |  |  |  |  |  |
| LUNA-04 | <20/P | * | <20/N | <20/N | <20/N | <20/P | <20/N | <20/P | <20/N | <20/P |
| LUNA-05 | <20/N | * | <20/P | <20/N | <20/N | <20/N | <20/N | <20/N | <20/P | <20/N |
| LUNA-10 | <20/N | * | <20/N | <20/N | <20/N | <20/P | <20/N | <20/P | <20/P | <20/N |
| LUNA-13 | <20/P | <20/N | <20/N | <20/N | <20/P | <20/N | <20/N | <20/N | <20/N | <20/P |
| LUNA-16 | <20/N | <20/N | <20/N | <20/P | <20/N | <20/N | <20/N | <20/N | 20 | <20/N |
| LUNA-22 | <20/N | <20/P | <20/N | <20/N | <20/N | <20/N | <20/P | <20/N | <20/N | <20/N |
| LUNA-24 | <20/P | <20/P | <20/P | <20/N | <20/N | <20/N | <20/N | <20/N | <20/N | <20/N |
| **PYR+VPA** |  |  |  |  |  |  |  |  |  |  |
| LUNA-06 | <20/N | * | <20/N | <20/N | <20/N | <20/N | <20/N | <20/N | <20/N | <20/N |
| LUNA-07 | <20/N | * | <20/N | <20/P | <20/N | <20/N | <20/N | <20/P | <20/P | <20/N |
| LUNA-12 | <20/N | * | <20/P | <20/N | <20/N | <20/N | <20/N | <20/N | <20/N | <20/N |
| LUNA-19 | <20/N | <20/N | <20/N | <20/N | <20/N | <20/N | <20/N | <20/N | <20/N | <20/P |
| LUNA-21 | <20/N | <20/N | <20/N | <20/N | <20/N | <20/N | <20/N | <20/N | <20/N | <20/N |
| LUNA-23 | <20/P | <20/N | <20/P | 22 | <20/P | <20/N | <20/P | <20/P | <20/N | <20/P |
| LUNA-25 | <20/N | <20/N | <20/P | <20/N | <20/P | <20/N | <20/P | <20/N | <20/N | <20/P |
| **VPA** |  |  |  |  |  |  |  |  |  |  |
| LUNA-02 | <20/N | * | <20/N | <20/N | <20/N | <20/N | <20/P | <20/N | <20/N | <20/N |
| LUNA-03 | <20/N | * | <20/N | <20/N | <20/N | <20/N | <20/N | <20/N | <20/N | <20/N |
| LUNA-09 | <20/N | * | <20/P | <20/N | <20/N | <20/N | <20/N | <20/N | <20/N | <20/P |
| LUNA-11 | <20/N | * | <20/N | <20/N | <20/N | <20/P | <20/N | <20/P | <20/N | <20/N |
| LUNA-18 | <20/N | <20/N | <20/N | <20/N | <20/N | <20/N | <20/N | <20/N | <20/N | <20/P |
| LUNA-20 | <20/P | <20/N | 41 | <20/P | <20/N | <20/P | <20/P | <20/P | <20/N | <20/P |
| LUNA-26 | <20/N | <20/P | <20/N | <20/N | <20/N | <20/N | <20/P | 21 | <20/N | <20/P |
| **CONTROL** |  |  |  |  |  |  |  |  |  |  |
| LUNA-01 | <20/N | * | <20/N | <20/N | <20/P | <20/N | <20/N | <20/N | <20/N | <20/N |
| LUNA-08 | <20/N | * | <20/N | <20/N | <20/P | <20/P | <20/N | <20/P | <20/P | <20/N |
| LUNA-14 | <20/N | <20/N | <20/N | <20/P | <20/N | <20/P | <20/N | <20/P | <20/P | <20/N |
| LUNA-15 | <20/N | <20/N | <20/P | <20/N | <20/N | <20/N | <20/N | <20/N | <20/N | <20/N |
| LUNA-17 | <20/N | <20/N | <20/N | <20/N | <20/N | <20/P | <20/N | <20/N | <20/N | <20/P |
| LUNA-27 | <20/N | <20/N | <20/N | <20/N | <20/N | <20/N | <20/N | <20/N | <20/N | <20/P |
| LUNA-28 | <20/N | <20/N | <20/P | <20/N | <20/P | <20/P | <20/N | <20/P | <20/N | <20/N |

\*Protocol violation where day 0 6 hour timepoint was not collected for participants LUNA-01 till LUNA-12.
/N denotes target viral genome not detected below level of quantitation.
/P denotes target viral genome detected below level of quantitation.
Those with and without pyrimethamine exposure were comparable with regard to the proportions of samples with plasma HIV-1 RNA above the assay detection limit both during routine care prior to study inclusion since cART initiation (20.7% versus 17.8%) and during the study at the fixed measurements (22.2% versus 23.6%).
Abbreviations: PYR: pyrimethamine; VPA: valproic acid.

**Table S7:**
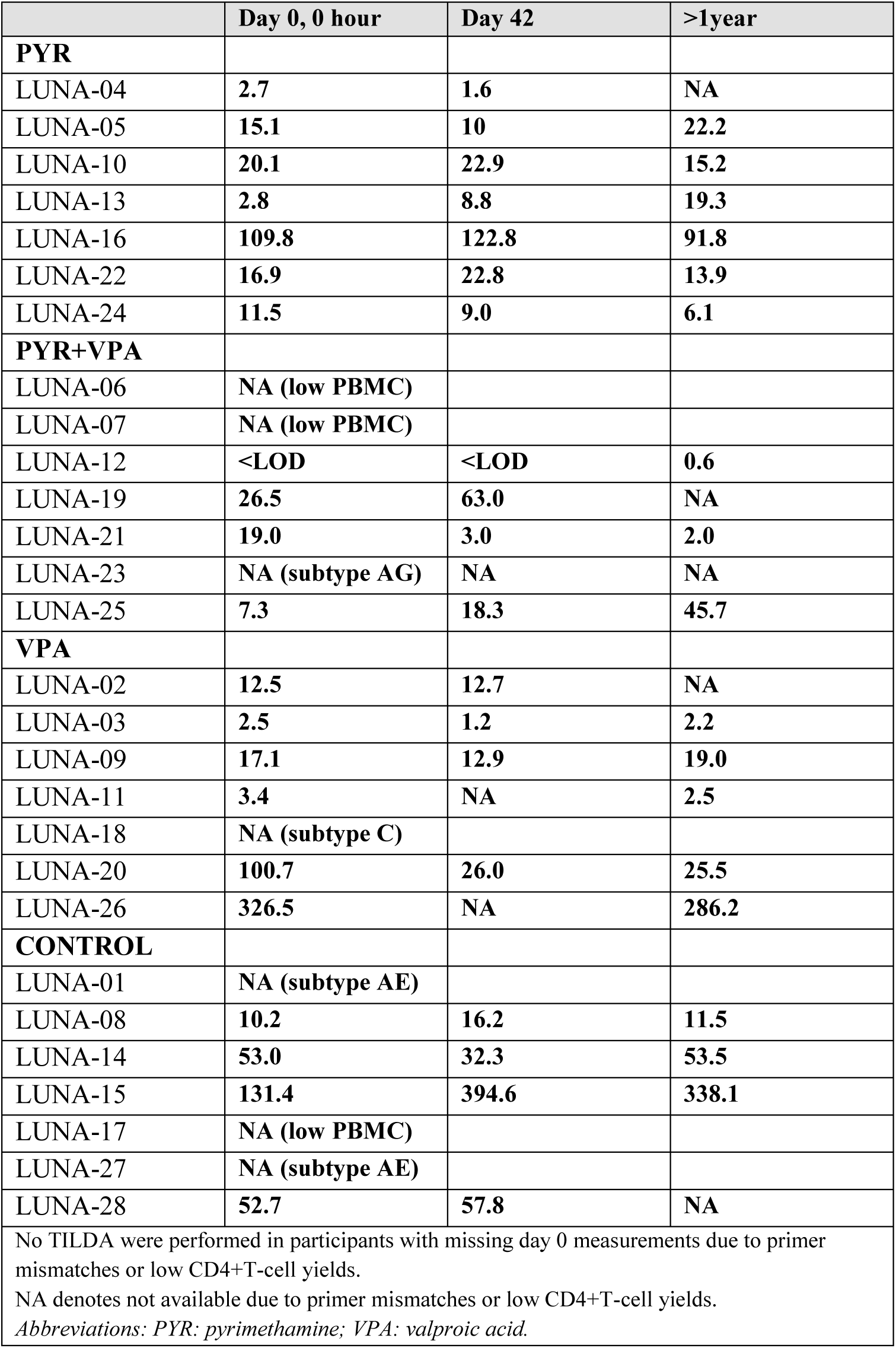
TILDA reservoir as number of cells with multiply spliced HIV-RNA detectable per million CD4+ T-cells per participant.

**Table S8:**
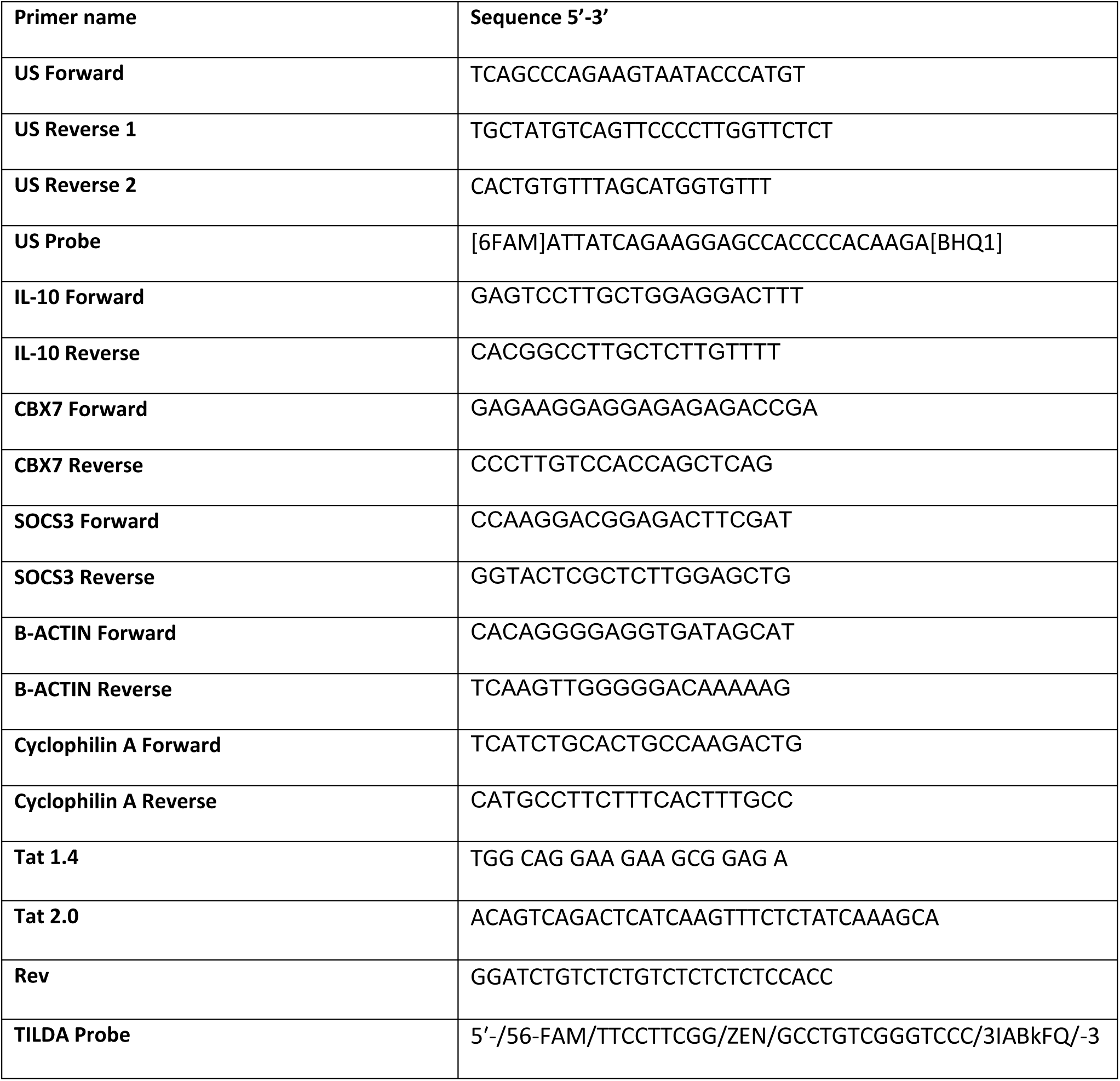
List of primers used for PCR for CA US HIV-1 RNA, TILDA and BAF complex target genes.

**Study protocol synopsis**

**Protocol version 3.0**

**27 January, 2020**

### Rationale

The retrovirus HIV integrates as proviral DNA in the genome of our CD4+ T cells. A subset forms a reservoir of latently infected long-lived memory T-cells with nearly absent HIV-DNA transcription. This persistent latent HIV reservoir is the major obstacle for a cure. HIV latency is sustained by multiple host factors that restrict the viral promotor and expression of the viral genome. Latency reversing agents (LRA) can remove these restrictive components and mediate HIV latency reversal. LRA monotherapy with histone deacetylase inhibitors (HDACi), including valproic acid, vorinostat, romidepsin, panobinostat, reactivates HIV but seems insufficient to eliminate the reservoir in vivo.

Our research group has identified the BAF complex as a repressive factor that maintains HIV latency. We investigated the activity of a panel of recently identified small molecule inhibitors of BAF (BAFi) as a new LRA group and showed that BAFi, including the clinically approved drug pyrimethamine at tolerable concentrations, are capable of reversing HIV latency and act synergistic with HDACi in vitro and in CD4+ T cells obtained from people with HIV on suppressive antiretroviral therapy. This offers new opportunities for cure research. We want to conduct the first study with BAFi and assess the potential synergism of 2 LRA with different modes of action on the reservoir in people with HIV.

### Design and primary objective

The LUNA study is a 6 week prospective, open label, randomized controlled clinical trial. The primary objective is the longitudinal assessment of the BAFi pyrimethamine and of the HDACi valproic acid on the HIV reservoir in people with HIV on antiretroviral therapy.

The main hypothesis tested is:

H0: the change in CA-US HIVRNA between the groups during treatment are equal.

H1: the change in CA-US HIVRNA between the groups during treatment are not equal.

### Study population

#### Inclusion criteria

1. HIV-1 infected patients ≥18 years.
2. WHO performance status 0 or 1.
3. Confirmed HIV-1 infection by 4th generation ELISA, Western Blot or PCR.
4. Wild type HIV infection or polymorphisms associated with at highest low-level resistance to any class of ART according to Stanford HIV drug resistance database. Transmitted mutations and acquired mutations due to virological failure associated with resistance of at highest low-level resistance are allowed.
5. On cART.
6. Current plasma HIV-RNA <50 copies/mL for at least 365 days and measured on at least 2 occasions of which at least 1 must be obtained within 365 and 90 days prior to study entry.
7. Current CD4 count at study entry of ≥200 cells/mm3.
8. Pre-cART HIV-RNA ≥10.000 copies/mL.

#### Exclusion criteria

A potential subject who meets any of the following criteria will be excluded from participation in this study.

1. Previous virological failure, defined as either acquired resistance mutations (>low level resistance) on cART or HIV-RNA >1000 copies/mL on two consecutive measurements during cART.
2. Uncontrolled hepatitis B or C co-infection.
3. Prior exposure to any HDACi, BAFi or other known LRA.
4. Prior exposure to cytotoxic myeloablative chemotherapy for hematological malignancies during cART.
5. Concurrent exposure to strong interacting medication on glucuronidation.
6. Exposure within 90 days prior to study entry to immunomodulators, cytokines, systemic antifungals, dexamethasone, vitamin K antagonists, anti-epileptics, antipsychotica, carbapenems, mefloquine, colestyramine, Any documented opportunistic infection related to HIV in the last 90 days.
7. Inadequate blood counts, renal and hepatic function tests.

a. Haemoglobin <6.5 mmol/L (males) or <6.0 mmol/L (females), leucocytes <2.5 x109/L, absolute neutrophil count <1000 cells/mm3, thrombocytes <100 x109/L, international standardized ratio >1.6, activated partial thromboplastin time >40 seconds.
b. Estimated glomerular filtration rate <50 mL/min (CKD-EPI).
c. ALAT or total bilirubin >2.5x upper limit of normal.
d. All laboratory values must be obtained within 42 days prior to the baseline visit.
8. Megaloblastic anemia due to folate deficiency.
9. Pancreatitis in last 6 months, or chronic pancreatitis.
10. Active malignancy during the past year with the exception of basal carcinoma of the skin, stage 0 cervical carcinoma, Kaposi Sarcoma treated with cART alone, or other indolent malignancies.
11. Females in the reproductive age cannot participate. Males cannot participate if they refuse to abstain from sex or condom use in serodiscordant sexual contact during the study, except if their sexual partner(s) use PREP.
12. Patients with active substance abuse or registered allergies to the investigational medical products.
13. Last, any other condition (familial, psychological, sociological, geographical) which in the investigator’s opinion poses an unacceptable risk or would hamper compliance with the study protocol and follow up schedule, will prohibit participation.

For hepatitis B: patients should be vaccinated, or on pre-exposure prophylaxis through the use of lamivudine/emtricitabine or tenofovir in their cART. Otherwise, standard serological testing should be available within the last 365 days for men with HIV who have sex with men. For other persons with HIV, there should be at least one negative hepatitis B test (either by serology or PCR) For men with HIV who have sex with men, a negative hepatitis C IgG, HCV antigen, blot or HCV-RNA PCR should be available within the previous 365 days. For other persons with HIV, there should be at least one negative hepatitis C test (either IgG, blot or PCR) available.

### Primary endpoint

The change in HIV reactivation in the reservoir in vivo at treatment initiation and at the end of treatment, measured as the change in cell associated HIV-RNA. The change in reactivation is compared between the treatment arms. The primary outcome measure is the change in cell associated HIV-RNA between treatment initiation (week 0) and at the end of study (week 6).

### Safety definitions

The study was terminated in case of excessive serious adverse events based on predefined safety criteria. Patients were monitored for adverse events. Study drugs were stopped if a drug-related SAE or AE of grade 4 or higher occurred, in case of AIDS-related illnesses CDC C, or if the blood CD4+ T cell count dropped below 200.

This was an open label study using approved drugs with a known safety profile for a shorter duration (pyrimethamine and valproic acid) than in usual care. The lab results (blood CD4+T-cells, plasma HIV RNA) and clinical condition that define the safety of this trial were readily available of all patients for the investigators. To protect the safety of the patients, the following study stopping rules were used:

1. A pre-specified interim analysis will be done after 14 patients (50%) are randomized. This will focus on the number of patients that had to discontinue treatment on investigators discretion due to blood CD4+T-cell count <200 during the IMP use, CDC-C events, possibly drug related SAE/AE ≥grade4, or AIDS related illness CDC C. If this exceeds 2 (10% of 21 patients that undergo an intervention) patients, the study will be stopped.
2. If ≥2 patients (of the 21 patients that undergo an intervention) have discontinued treatment due to possibly drug related AE before the planned interim analysis is done, the study will be stopped.
3. If at any time in the study the number of discontinuations due to blood CD4+T-cell count <200 during IMP use, or CDC-C events or possibly drug related SAE/AE ≥grade4, AIDS related illness CDC C exceeds 25% of 21 patients undergoing an intervention (>5 patients), the study will be stopped.
4. If ≥2 patients (10%) experience HIV treatment failure with acquisition of resistance associated mutations in HIV, the study will be stopped.

## REFERENCES

1. T. W. Chun, D. Engel, M. M. Berrey, T. Shea, L. Corey, A. S. Fauci, Early establishment of a pool of latently infected, resting CD4(+) T cells during primary HIV-1 infection. Proc Natl Acad Sci U S A 95, 8869–8873 (1998).

2. T. W. Chun, R. T. Davey, Jr., M. Ostrowski, J. Shawn Justement, D. Engel, J. I. Mullins, A. S. Fauci, Relationship between pre-existing viral reservoirs and the re-emergence of plasma viremia after discontinuation of highly active anti-retroviral therapy. Nat Med 6, 757–761 (2000).

3. G. Strategies for Management of Antiretroviral Therapy Study, W. M. El-Sadr, J. Lundgren, J. D. Neaton, F. Gordin, D. Abrams, R. C. Arduino, A. Babiker, W. Burman, N. Clumeck, C. J. Cohen, D. Cohn, D. Cooper, J. Darbyshire, S. Emery, G. Fatkenheuer, B. Gazzard, B. Grund, J. Hoy, K. Klingman, M. Losso, N. Markowitz, J. Neuhaus, A. Phillips, C. Rappoport, CD4+ count-guided interruption of antiretroviral treatment. N Engl J Med 355, 2283–2296 (2006).

4. S. G. Deeks, HIV: Shock and kill. Nature 487, 439–440 (2012).

5. S. G. Deeks, N. Archin, P. Cannon, S. Collins, R. B. Jones, M. de Jong, O. Lambotte, R. Lamplough, T. Ndung’u, J. Sugarman, C. T. Tiemessen, L. Vandekerckhove, S. R. Lewin, A. S. G. S. S. w. g. International, Research priorities for an HIV cure: International AIDS Society Global Scientific Strategy 2021. Nat Med 27, 2085–2098 (2021).

6. D. M. Margolis, N. M. Archin, M. S. Cohen, J. J. Eron, G. Ferrari, J. V. Garcia, C. L. Gay, N. Goonetilleke, S. B. Joseph, R. Swanstrom, A. W. Turner, A. Wahl, Curing HIV: Seeking to Target and Clear Persistent Infection. Cell 181, 189–206 (2020).

7. M. Stoszko, E. Ne, E. Abner, T. Mahmoudi, A broad drug arsenal to attack a strenuous latent HIV reservoir. Curr Opin Virol 38, 37–53 (2019).

8. A. Ait-Ammar, A. Kula, G. Darcis, R. Verdikt, S. De Wit, V. Gautier, P. W. G. Mallon, A. Marcello, O. Rohr, C. Van Lint, Current Status of Latency Reversing Agents Facing the Heterogeneity of HIV-1 Cellular and Tissue Reservoirs. Front Microbiol 10, 3060 (2019).

9. S. A. Yukl, P. Kaiser, P. Kim, S. Telwatte, S. K. Joshi, M. Vu, H. Lampiris, J. K. Wong, HIV latency in isolated patient CD4(+) T cells may be due to blocks in HIV transcriptional elongation, completion, and splicing. Sci Transl Med 10, (2018).

10. S. Telwatte, S. Moron-Lopez, D. Aran, P. Kim, C. Hsieh, S. Joshi, M. Montano, W. C. Greene, A. J. Butte, J. K. Wong, S. A. Yukl, Heterogeneity in HIV and cellular transcription profiles in cell line models of latent and productive infection: implications for HIV latency. Retrovirology 16, 32 (2019).

11. S. Rao, C. Lungu, R. Crespo, T. H. Steijaert, A. Gorska, R. J. Palstra, H. A. B. Prins, W. van Ijcken, Y. M. Mueller, J. J. A. van Kampen, A. Verbon, P. D. Katsikis, C. A. B. Boucher, C. Rokx, R. A. Gruters, T. Mahmoudi, Selective cell death in HIV-1-infected cells by DDX3 inhibitors leads to depletion of the inducible reservoir. Nat Commun 12, 2475 (2021).

12. R. Crespo, S. Rao, T. Mahmoudi, HibeRNAtion: HIV-1 RNA Metabolism and Viral Latency. Front Cell Infect Microbiol 12, 855092 (2022).

13. A. Kumar, G. Darcis, C. Van Lint, G. Herbein, Epigenetic control of HIV-1 post integration latency: implications for therapy. Clin Epigenetics 7, 103 (2015).

14. C. Van Lint, S. Bouchat, A. Marcello, HIV-1 transcription and latency: an update. Retrovirology 10, 67 (2013).

15. E. Ne, R. J. Palstra, T. Mahmoudi, Transcription: Insights From the HIV-1 Promoter. Int Rev Cell Mol Biol 335, 191–243 (2018).

16. F. Wightman, P. Ellenberg, M. Churchill, S. R. Lewin, HDAC inhibitors in HIV. Immunol Cell Biol 90, 47–54 (2012).

17. G. Lehrman, I. B. Hogue, S. Palmer, C. Jennings, C. A. Spina, A. Wiegand, A. L. Landay, R. W. Coombs, D. D. Richman, J. W. Mellors, J. M. Coffin, R. J. Bosch, D. M. Margolis, Depletion of latent HIV-1 infection in vivo: a proof-of-concept study. Lancet 366, 549–555 (2005).

18. J. D. Siliciano, J. Lai, M. Callender, E. Pitt, H. Zhang, J. B. Margolick, J. E. Gallant, J. Cofrancesco, Jr., R. D. Moore, S. J. Gange, R. F. Siliciano, Stability of the latent reservoir for HIV-1 in patients receiving valproic acid. J Infect Dis 195, 833–836 (2007).

19. N. M. Archin, J. J. Eron, S. Palmer, A. Hartmann-Duff, J. A. Martinson, A. Wiegand, N. Bandarenko, J. L. Schmitz, R. J. Bosch, A. L. Landay, J. M. Coffin, D. M. Margolis, Valproic acid without intensified antiviral therapy has limited impact on persistent HIV infection of resting CD4+ T cells. AIDS 22, 1131–1135 (2008).

20. N. Sagot-Lerolle, A. Lamine, M. L. Chaix, F. Boufassa, J. P. Aboulker, D. Costagliola, C. Goujard, C. Pallier, J. F. Delfraissy, O. Lambotte, A. E. study, Prolonged valproic acid treatment does not reduce the size of latent HIV reservoir. AIDS 22, 1125–1129 (2008).

21. N. M. Archin, M. Cheema, D. Parker, A. Wiegand, R. J. Bosch, J. M. Coffin, J. Eron, M. Cohen, D. M. Margolis, Antiretroviral intensification and valproic acid lack sustained effect on residual HIV-1 viremia or resting CD4+ cell infection. PLoS One 5, e9390 (2010).

22. J. P. Routy, C. L. Tremblay, J. B. Angel, B. Trottier, D. Rouleau, J. G. Baril, M. Harris, S. Trottier, J. Singer, N. Chomont, R. P. Sekaly, M. R. Boulassel, Valproic acid in association with highly active antiretroviral therapy for reducing systemic HIV-1 reservoirs: results from a multicentre randomized clinical study. HIV Med 13, 291–296 (2012).

23. N. M. Archin, A. Espeseth, D. Parker, M. Cheema, D. Hazuda, D. M. Margolis, Expression of latent HIV induced by the potent HDAC inhibitor suberoylanilide hydroxamic acid. AIDS Res Hum Retroviruses 25, 207–212 (2009).

24. N. M. Archin, A. L. Liberty, A. D. Kashuba, S. K. Choudhary, J. D. Kuruc, A. M. Crooks, D. C. Parker, E. M. Anderson, M. F. Kearney, M. C. Strain, D. D. Richman, M. G. Hudgens, R. J. Bosch, J. M. Coffin, J. J. Eron, D. J. Hazuda, D. M. Margolis, Administration of vorinostat disrupts HIV-1 latency in patients on antiretroviral therapy. Nature 487, 482–485 (2012).

25. N. M. Archin, R. Bateson, M. K. Tripathy, A. M. Crooks, K. H. Yang, N. P. Dahl, M. F. Kearney, E. M. Anderson, J. M. Coffin, M. C. Strain, D. D. Richman, K. R. Robertson, A. D. Kashuba, R. J. Bosch, D. J. Hazuda, J. D. Kuruc, J. J. Eron, D. M. Margolis, HIV-1 expression within resting CD4+ T cells after multiple doses of vorinostat. J Infect Dis 210, 728–735 (2014).

26. J. H. Elliott, F. Wightman, A. Solomon, K. Ghneim, J. Ahlers, M. J. Cameron, M. Z. Smith, T. Spelman, J. McMahon, P. Velayudham, G. Brown, J. Roney, J. Watson, M. H. Prince, J. F. Hoy, N. Chomont, R. Fromentin, F. A. Procopio, J. Zeidan, S. Palmer, L. Odevall, R. W. Johnstone, B. P. Martin, E. Sinclair, S. G. Deeks, D. J. Hazuda, P. U. Cameron, R. P. Sekaly, S. R. Lewin, Activation of HIV transcription with short-course vorinostat in HIV-infected patients on suppressive antiretroviral therapy. PLoS Pathog 10, e1004473 (2014).

27. N. M. Archin, J. L. Kirchherr, J. A. Sung, G. Clutton, K. Sholtis, Y. Xu, B. Allard, E. Stuelke, A. D. Kashuba, J. D. Kuruc, J. Eron, C. L. Gay, N. Goonetilleke, D. M. Margolis, Interval dosing with the HDAC inhibitor vorinostat effectively reverses HIV latency. J Clin Invest 127, 3126–3135 (2017).

28. S. Fidler, W. Stohr, M. Pace, L. Dorrell, A. Lever, S. Pett, S. Kinloch-de Loes, J. Fox, A. Clarke, M. Nelson, J. Thornhill, M. Khan, A. Fun, M. Bandara, D. Kelly, J. Kopycinski, T. Hanke, H. Yang, R. Bennett, M. Johnson, B. Howell, R. Barnard, G. Wu, S. Kaye, M. Wills, A. Babiker, J. Frater, R. t. s. group, Antiretroviral therapy alone versus antiretroviral therapy with a kick and kill approach, on measures of the HIV reservoir in participants with recent HIV infection (the RIVER trial): a phase 2, randomised trial. Lancet 395, 888–898 (2020).

29. J. C. Ramos, J. A. Sparano, A. Chadburn, E. G. Reid, R. F. Ambinder, E. R. Siegel, P. C. Moore, P. G. Rubinstein, C. M. Durand, E. Cesarman, D. Aboulafia, R. Baiocchi, L. Ratner, L. Kaplan, A. A. Capoferri, J. Y. Lee, R. Mitsuyasu, A. Noy, Impact of Myc in HIV-associated non-Hodgkin lymphomas treated with EPOCH and outcomes with vorinostat (AMC-075 trial). Blood 136, 1284–1297 (2020).

30. E. Kroon, J. Ananworanich, A. Pagliuzza, A. Rhodes, N. Phanuphak, L. Trautmann, J. L. Mitchell, M. Chintanaphol, J. Intasan, S. Pinyakorn, K. Benjapornpong, J. J. Chang, D. J. Colby, N. Chomchey, J. L. K. Fletcher, K. Eubanks, H. Yang, J. Kapson, A. Dantanarayana, S. Tennakoon, R. J. Gorelick, F. Maldarelli, M. L. Robb, J. H. Kim, S. Spudich, N. Chomont, P. Phanuphak, S. R. Lewin, M. S. de Souza, Search, R. V. S. Teams, A randomized trial of vorinostat with treatment interruption after initiating antiretroviral therapy during acute HIV-1 infection. J Virus Erad 6, 100004 (2020).

31. C. L. Gay, K. S. James, M. Tuyishime, S. D. Falcinelli, S. B. Joseph, M. J. Moeser, B. Allard, J. L. Kirchherr, M. Clohosey, S. L. M. Raines, D. C. Montefiori, X. Shen, R. J. Gorelick, L. Gama, A. B. McDermott, R. A. Koup, J. R. Mascola, M. Floris-Moore, J. D. Kuruc, G. Ferrari, J. J. Eron, N. M. Archin, D. M. Margolis, Stable Latent HIV Infection and Low-level Viremia Despite Treatment With the Broadly Neutralizing Antibody VRC07-523LS and the Latency Reversal Agent Vorinostat. J Infect Dis 225, 856–861 (2022).

32. J. H. McMahon, V. A. Evans, J. S. Y. Lau, J. Symons, J. M. Zerbato, J. Chang, A. Solomon, S. Tennakoon, A. Dantanarayana, M. Hagenauer, S. Lee, S. Palmer, K. Fisher, N. Bumpus, C. J. S. Heck, D. Burger, G. Wu, P. Zuck, B. J. Howell, H. H. Zetterberg, K. Blennow, M. Gisslen, T. A. Rasmussen, S. R. Lewin, Neurotoxicity with high-dose disulfiram and vorinostat used for HIV latency reversal. AIDS 36, 75–82 (2022).

33. A. E. P. Scully, E. Aga, A. Tsibris, N. Archin, K. Starr, Q. Ma, G. D. Morse, K. E. Squires, B. J. Howell, G. Wu, L. Hosey, S. Sieg, L. Ehui, F. Giguel, K. Coxen, C. Dobrowlski, M. Gandhi, S. Deeks, N. Chomont, E. Connick, C. Godfrey, J. Karn, D. R. Kuritzkes, R. J. Bosch, R. T. Gandhi, A. s. team, Impact of Tamoxifen on Vorinostat-Induced HIV Expression in Women on Antiretroviral Therapy (ACTG A5366:The MOXIE trial). Clin Infect Dis, (2022).

34. T. A. Rasmussen, M. Tolstrup, C. R. Brinkmann, R. Olesen, C. Erikstrup, A. Solomon, A. Winckelmann, S. Palmer, C. Dinarello, M. Buzon, M. Lichterfeld, S. R. Lewin, L. Ostergaard, O. S. Sogaard, Panobinostat, a histone deacetylase inhibitor, for latent-virus reactivation in HIV-infected patients on suppressive antiretroviral therapy: a phase 1/2, single group, clinical trial. Lancet HIV 1, e13–21 (2014).

35. O. S. Sogaard, M. E. Graversen, S. Leth, R. Olesen, C. R. Brinkmann, S. K. Nissen, A. S. Kjaer, M. H. Schleimann, P. W. Denton, W. J. Hey-Cunningham, K. K. Koelsch, G. Pantaleo, K. Krogsgaard, M. Sommerfelt, R. Fromentin, N. Chomont, T. A. Rasmussen, L. Ostergaard, M. Tolstrup, The Depsipeptide Romidepsin Reverses HIV-1 Latency In Vivo. PLoS Pathog 11, e1005142 (2015).

36. A. S. Leth, M. H. Schleimann, S. K. Nissen, J. F. Hojen, R. Olesen, M. E. Graversen, S. Jorgensen, A. S. Kjaer, P. W. Denton, A. Mork, M. A. Sommerfelt, K. Krogsgaard, L. Ostergaard, T. A. Rasmussen, M. Tolstrup, O. S. Sogaard, Combined effect of Vacc-4x, recombinant human granulocyte macrophage colony-stimulating factor vaccination, and romidepsin on the HIV-1 reservoir (REDUC): a single-arm, phase 1B/2A trial. Lancet HIV 3, e463–472 (2016).

37. B. Mothe, M. Rosas-Umbert, P. Coll, C. Manzardo, M. C. Puertas, S. Moron-Lopez, A. Llano, C. Miranda, S. Cedeno, M. Lopez, Y. Alarcon-Soto, G. G. Melis, K. Langohr, A. M. Barriocanal, J. Toro, I. Ruiz, C. Rovira, A. Carrillo, M. Meulbroek, A. Crook, E. G. Wee, J. M. Miro, B. Clotet, M. Valle, J. Martinez-Picado, T. Hanke, C. Brander, J. Molto, B. C. N. S. Investigators, HIVconsv Vaccines and Romidepsin in Early-Treated HIV-1-Infected Individuals: Safety, Immunogenicity and Effect on the Viral Reservoir (Study BCN02). Front Immunol 11, 823 (2020).

38. D. K. McMahon, L. Zheng, J. C. Cyktor, E. Aga, B. J. Macatangay, C. Godfrey, M. Para, R. T. Mitsuyasu, J. Hesselgesser, J. Dragavon, C. Dobrowolski, J. Karn, E. P. Acosta, R. T. Gandhi, J. W. Mellors, A Phase 1/2 Randomized, Placebo-Controlled Trial of Romidespin in Persons With HIV-1 on Suppressive Antiretroviral Therapy. J Infect Dis 224, 648–656 (2021).

39. H. Gruell, J. D. Gunst, Y. Z. Cohen, M. H. Pahus, J. J. Malin, M. Platten, K. G. Millard, M. Tolstrup, R. B. Jones, W. D. Conce Alberto, J. C. C. Lorenzi, T. Y. Oliveira, T. Kummerle, I. Suarez, C. Unson-O’Brien, L. Nogueira, R. Olesen, L. Ostergaard, H. Nielsen, C. Lehmann, M. C. Nussenzweig, G. Fatkenheuer, F. Klein, M. Caskey, O. S. Sogaard, Effect of 3BNC117 and romidepsin on the HIV-1 reservoir in people taking suppressive antiretroviral therapy (ROADMAP): a randomised, open-label, phase 2A trial. Lancet Microbe 3, e203–e214 (2022).

40. J. H. Elliott, J. H. McMahon, C. C. Chang, S. A. Lee, W. Hartogensis, N. Bumpus, R. Savic, J. Roney, R. Hoh, A. Solomon, M. Piatak, R. J. Gorelick, J. Lifson, P. Bacchetti, S. G. Deeks, S. R. Lewin, Short-term administration of disulfiram for reversal of latent HIV infection: a phase 2 dose-escalation study. Lancet HIV 2, e520–529 (2015).

41. C. Gutierrez, S. Serrano-Villar, N. Madrid-Elena, M. J. Perez-Elias, M. E. Martin, C. Barbas, J. Ruiperez, E. Munoz, M. A. Munoz-Fernandez, T. Castor, S. Moreno, Bryostatin-1 for latent virus reactivation in HIV-infected patients on antiretroviral therapy. AIDS 30, 1385–1392 (2016).

42. A. L. Vibholm, M. H. Schleimann, J. F. Hojen, T. Benfield, R. Offersen, K. Rasmussen, R. Olesen, A. Dige, J. Agnholt, J. Grau, M. Buzon, B. Wittig, M. Lichterfeld, A. M. Petersen, X. Deng, M. Abdel-Mohsen, S. K. Pillai, S. Rutsaert, W. Trypsteen, W. De Spiegelaere, L. Vandekerchove, L. Ostergaard, T. A. Rasmussen, P. W. Denton, M. Tolstrup, O. S. Sogaard, Short-Course Toll-Like Receptor 9 Agonist Treatment Impacts Innate Immunity and Plasma Viremia in Individuals With Human Immunodeficiency Virus Infection. Clin Infect Dis 64, 1686–1695 (2017).

43. T. S. Uldrick, S. V. Adams, R. Fromentin, M. Roche, S. P. Fling, P. H. Goncalves, K. Lurain, R. Ramaswami, C. J. Wang, R. J. Gorelick, J. L. Welker, L. O’Donoghue, H. Choudhary, J. D. Lifson, T. A. Rasmussen, A. Rhodes, C. Tumpach, R. Yarchoan, F. Maldarelli, M. A. Cheever, R. Sekaly, N. Chomont, S. G. Deeks, S. R. Lewin, Pembrolizumab induces HIV latency reversal in people living with HIV and cancer on antiretroviral therapy. Sci Transl Med 14, eabl3836 (2022).

44. A. M. Stoszko, E. De Crignis, C. Rokx, M. M. Khalid, C. Lungu, R. J. Palstra, T. W. Kan, C. Boucher, A. Verbon, E. C. Dykhuizen, T. Mahmoudi, Small Molecule Inhibitors of BAF; A Promising Family of Compounds in HIV-1 Latency Reversal. EBioMedicine 3, 108–121 (2016).

45. C. A. Marian, M. Stoszko, L. Wang, M. W. Leighty, E. de Crignis, C. A. Maschinot, J. Gatchalian, B. C. Carter, B. Chowdhury, D. C. Hargreaves, J. R. Duvall, G. R. Crabtree, T. Mahmoudi, E. C. Dykhuizen, Small Molecule Targeting of Specific BAF (mSWI/SNF) Complexes for HIV Latency Reversal. Cell Chem Biol 25, 1443–1455 e1414 (2018).

46. E. Ne, R. Crespo, R. Izquierdo-Lara, S. Rao, S. Kocer, A. Gorska, T. van Staveren, T. W. Kan, D. van de Vijver, D. Dekkers, C. Rokx, P. Moulos, P. Hatzis, R. J. Palstra, J. Demmers, T. Mahmoudi, Catchet-MS identifies IKZF1-targeting thalidomide analogues as novel HIV-1 latency reversal agents. Nucleic Acids Res 50, 5577–5598 (2022).

47. T. Mahmoudi, The BAF complex and HIV latency. Transcription 3, 171–176 (2012).

48. H. Rafati, M. Parra, S. Hakre, Y. Moshkin, E. Verdin, T. Mahmoudi, Repressive LTR nucleosome positioning by the BAF complex is required for HIV latency. PLoS Biol 9, e1001206 (2011).

49. J. M. Jacobson, M. Davidian, P. M. Rainey, R. Hafner, R. H. Raasch, B. J. Luft, Pyrimethamine pharmacokinetics in human immunodeficiency virus-positive patients seropositive for Toxoplasma gondii. Antimicrob Agents Chemother 40, 1360–1365 (1996).

50. H. Klinker, P. Langmann, E. Richter, Pyrimethamine alone as prophylaxis for cerebral toxoplasmosis in patients with advanced HIV infection. Infection 24, 324–327 (1996).

51. H. Klinker, P. Langmann, E. Richter, Plasma pyrimethamine concentrations during long-term treatment for cerebral toxoplasmosis in patients with AIDS. Antimicrob Agents Chemother 40, 1623–1627 (1996).

52. D. R. Schmidt, B. Hogh, O. Andersen, S. H. Hansen, K. Dalhoff, E. Petersen, Treatment of infants with congenital toxoplasmosis: tolerability and plasma concentrations of sulfadiazine and pyrimethamine. Eur J Pediatr 165, 19–25 (2006).

53. S. Kongsaengdao, K. Samintarapanya, K. Oranratnachai, W. Prapakarn, C. Apichartpiyakul, Randomized controlled trial of pyrimethamine plus sulfadiazine versus trimethoprim plus sulfamethoxazole for treatment of toxoplasmic encephalitis in AIDS patients. J Int Assoc Physicians AIDS Care (Chic) 7, 11–16 (2008).

54. J. M. Jacobson, R. Hafner, J. Remington, C. Farthing, J. Holden-Wiltse, E. M. Bosler, C. Harris, D. T. Jayaweera, C. Roque, B. J. Luft, A. S. Team, Dose-escalation, phase I/II study of azithromycin and pyrimethamine for the treatment of toxoplasmic encephalitis in AIDS. AIDS 15, 583–589 (2001).

55. H. W. Unger, M. Ome-Kaius, R. A. Wangnapi, A. J. Umbers, S. Hanieh, C. S. Suen, L. J. Robinson, A. Rosanas-Urgell, J. Wapling, E. Lufele, C. Kongs, P. Samol, D. Sui, D. Singirok, A. Bardaji, L. Schofield, C. Menendez, I. Betuela, P. Siba, I. Mueller, S. J. Rogerson, Sulphadoxine-pyrimethamine plus azithromycin for the prevention of low birthweight in Papua New Guinea: a randomised controlled trial. BMC Med 13, 9 (2015).

56. K. Chirgwin, R. Hafner, C. Leport, J. Remington, J. Andersen, E. M. Bosler, C. Roque, N. Rajicic, V. McAuliffe, P. Morlat, D. T. Jayaweera, J. L. Vilde, B. J. Luft, Randomized phase II trial of atovaquone with pyrimethamine or sulfadiazine for treatment of toxoplasmic encephalitis in patients with acquired immunodeficiency syndrome: ACTG 237/ANRS 039 Study. AIDS Clinical Trials Group 237/Agence Nationale de Recherche sur le SIDA, Essai 039. Clin Infect Dis 34, 1243-1250 (2002).

57. US Food & Drug Administration. Pyrimethamine Summary of Product Charachteristics. Available at: http://www.accessdata.fda.gov/drugsatfda_docs/label/2003/08578slr016_daraprim_lbl.pdf.

58. R. McLeod, D. Mack, R. Foss, K. Boyer, S. Withers, S. Levin, J. Hubbell, Levels of pyrimethamine in sera and cerebrospinal and ventricular fluids from infants treated for congenital toxoplasmosis. Toxoplasmosis Study Group. Antimicrob Agents Chemother 36, 1040–1048 (1992).

59. P. D. J. Bollen, H. A. B. Prins, A. Colbers, K. Velthoven-Graafland, B. J. A. Rijnders, T. de Vries-Sluijs, E. van Nood, J. Nouwen, H. Bax, M. de Mendonca Melo, A. Verbon, D. M. Burger, C. Rokx, The dolutegravir/valproic acid drug-drug interaction is primarily based on protein displacement. J Antimicrob Chemother 76, 1273–1276 (2021).

60. US Food & Drug Administration. Valproic Acid Summary of Product Characteristics. Available at: https://www.accessdata.fda.gov/drugsatfda_docs/label/2009/018081s047,018082s032lbl.pdf.

61. A. L. Wurster, P. Precht, K. G. Becker, W. H. Wood, 3rd, Y. Zhang, Z. Wang, M. J. Pazin, IL-10 transcription is negatively regulated by BAF180, a component of the SWI/SNF chromatin remodeling enzyme. BMC Immunol 13, 9 (2012).

62. E. C. Dykhuizen, L. C. Carmody, N. Tolliday, G. R. Crabtree, M. A. Palmer, Screening for inhibitors of an essential chromatin remodeler in mouse embryonic stem cells by monitoring transcriptional regulation. J Biomol Screen 17, 1221–1230 (2012).

63. N. Barker, A. Hurlstone, H. Musisi, A. Miles, M. Bienz, H. Clevers, The chromatin remodelling factor Brg-1 interacts with beta-catenin to promote target gene activation. EMBO J 20, 4935–4943 (2001).

64. Z. Ni, R. Bremner, Brahma-related gene 1-dependent STAT3 recruitment at IL-6-inducible genes. J Immunol 178, 345–351 (2007).

65. F. A. Procopio, R. Fromentin, D. A. Kulpa, J. H. Brehm, A. G. Bebin, M. C. Strain, D. D. Richman, U. O’Doherty, S. Palmer, F. M. Hecht, R. Hoh, R. J. Barnard, M. D. Miller, D. J. Hazuda, S. G. Deeks, R. P. Sekaly, N. Chomont, A Novel Assay to Measure the Magnitude of the Inducible Viral Reservoir in HIV-infected Individuals. EBioMedicine 2, 874–883 (2015).

66. C. Lungu, F. A. Procopio, R. J. Overmars, R. J. J. Beerkens, J. J. C. Voermans, S. Rao, H. A. B. Prins, C. Rokx, G. Pantaleo, D. Vijver, T. Mahmoudi, C. A. B. Boucher, R. A. Gruters, J. Kampen, Inter-Laboratory Reproducibility of Inducible HIV-1 Reservoir Quantification by TILDA. Viruses 12, (2020).

67. S. Y. Kim, R. Byrn, J. Groopman, D. Baltimore, Temporal aspects of DNA and RNA synthesis during human immunodeficiency virus infection: evidence for differential gene expression. J Virol 63, 3708–3713 (1989).

68. C. Mary, J. N. Telles, V. Cheynet, G. Oriol, F. Mallet, B. Mandrand, B. Verrier, Quantitative and discriminative detection of individual HIV-1 mRNA subspecies by an RNAse mapping assay. J Virol Methods 49, 9–23 (1994).

69. C. A. van Baalen, C. Guillon, M. van Baalen, E. J. Verschuren, P. H. Boers, A. D. Osterhaus, R. A. Gruters, Impact of antigen expression kinetics on the effectiveness of HIV-specific cytotoxic T lymphocytes. Eur J Immunol 32, 2644–2652 (2002).

70. A. O. Pasternak, K. W. Adema, M. Bakker, S. Jurriaans, B. Berkhout, M. Cornelissen, V. V. Lukashov, Highly sensitive methods based on seminested real-time reverse transcription-PCR for quantitation of human immunodeficiency virus type 1 unspliced and multiply spliced RNA and proviral DNA. J Clin Microbiol 46, 2206–2211 (2008).

71. T. D. Schmittgen, K. J. Livak, Analyzing real-time PCR data by the comparative C(T) method. Nat Protoc 3, 1101–1108 (2008).

